# Inferring time-varying generation time, serial interval and incubation period distributions for COVID-19

**DOI:** 10.1101/2022.08.05.22278461

**Authors:** Dongxuan Chen, Yiu Chung Lau, Xiao-Ke Xu, Lin Wang, Zhanwei Du, Tim K. Tsang, Peng Wu, Eric H. Y. Lau, Jacco Wallinga, Benjamin J. Cowling, Sheikh Taslim Ali

## Abstract

**Background:** The generation time distribution, reflecting the time between successive infections in transmission chains, is one of the fundamental epidemiological parameters for describing COVID-19 transmission dynamics. However, because exact infection times are rarely known, it is often approximated by the serial interval distribution, reflecting the time between illness onsets of infector and infectee. This approximation holds under the assumption that infectors and infectees share the same incubation period distribution, which may not always be true.

**Methods:** We analyzed data on observed incubation period and serial interval distributions in China, during January and February 2020, under different sampling approaches, and developed an inferential framework to estimate the generation time distribution that accounts for variation over time due to changes in epidemiology, sampling biases and public health and social measures.

**Results:** We analyzed data on a total of 2989 confirmed cases for COVID-19 during January 1 to February 29, 2020 in Mainland China. During the study period, the empirical forward serial interval decreased from a mean of 8.90 days to 2.68 days. The estimated mean backward incubation period of infectors increased from 3.77 days to 9.61 days, and the mean forward incubation period of infectees also increased from 5.39 days to 7.21 days. The estimated mean forward generation time decreased from 7.27 days (95% confidence interval: 6.42, 8.07) to 4.21 days (95% confidence interval: 3.70, 4.74) days by January 29. We used simulations to examine the sensitivity of our modelling approach to a number of assumptions and alternative dynamics.

**Conclusions:** The proposed method can provide more reliable estimation of the temporal variation in the generation time distribution, enabling proper assessment of transmission dynamics.

## Introduction

The coronavirus disease 2019 (COVID-19) pandemic has caused over 557 million cases and 6 million deaths by July 15, 2022^1^. The generation time (GT) distribution is one of the key transmission parameters and defined as the time between successive infections in a transmission chain. The generation time distribution shapes the relationship between epidemic growth rate and reproduction number^2^, while the reproduction number has been widely used to indicate the measure of transmissibility, and is defined as the average number of secondary cases infected by one typical infector in the population.

Exact infection times are hard to observe, hence the generation time distribution is usually unobserved. It is easier to record symptom onset times. Thus, in practice, the time between the illness onsets of infector and infectee, which is called the serial interval (SI), is commonly used as a proxy for the GT. Under the assumption that the infector and infectee have the same incubation period (IP) distribution, the mean SI would equal the mean GT^3,4^. Therefore, the entire serial interval distribution is often used to estimate the reproduction number^5,6^. However, this parametric approximation does not always hold, as GT and SI have different distributional properties. Importantly, the SI can be negative when the infectee has onset earlier than infector as shown in the pre-symptomatic transmission for COVID-19^7,8^, while GT must be positive since the infectee’s infection time must be later than infector’s infection time. In addition, the SI always has a larger variance than GT due to their different biological and clinical characteristics^9^. Thus when mean SI equals mean GT, using SI distribution as a proxy of GT distribution may underestimate the reproduction number^10–12^.

Sampling biases can also affect the estimation of transmission parameters^10,11^. While following up the cases since their infection time (i.e. forward sampling) would result in correct estimation of IP, case sampling with reference to onset times (i.e. backward sampling) would favour underestimation and overestimation of IP during the exponential and fading phase of the epidemic respectively^10^. Sampling with reference to infectee onset times, regarded as backward sampling of SI, will have the same issue. Moreover, sampling with reference to infector onset times, regarded as forward sampling of SI, also results in time-varying estimates of SI, as Park et al^11^ showed that the forward SI can be decomposed as the forward GT plus the forward IP of infectee minus the backward IP of infector, and posited that the decreasing trend of forward SI over time was due to the overestimation of infector’s IP under the backward sampling approach. Therefore, it is not appropriate to directly use temporal forward SI as a proxy of temporal GT. Following Park’s hypotheses, in this study, we developed an inferential framework to estimate the time-varying forward GT, hence to have more accurate estimation of the reproduction number. We apply this framework to observations on IP and SI in China during the first months of the COVID-19 pandemic, and quantify the actual magnitude of temporal variations in the estimates and their impact on the estimated generation times and reproduction numbers.

## Results

### Construction of transmission pairs

We investigated a total of 2989 confirmed cases for COVID-19 during January 1 to February 29, 2020 in Mainland China. Of these 2989 cases, the median age was 46 years-old (interquartile range (IQR): 33 – 58), and the proportion of male and female was 51% and 49% respectively. We reconstructed 629 transmission pairs having symptom onset times for both infectors and infectees, which consisted of 428 infectors and 629 infectees. Among the 428 infectors, the median age was 47 years-old (IQR: 37 – 57), and 59% were male; while among the 629 infectees, the median age was 49 years-old (IQR: 34 – 61), and 47% were male. The mean number of infectees infected by an infector in our data was 1.47, 386 (90%) infectors had no more than 2 infectees, while 4 (1%) infectors had more than 5 infectees, with the maximum of 16 infectees being suspected to have been infected by one single infector.

Despite the unknown infection times, the incubation period could be inferred by onset time and the exposure window as from the first to the last day of the case’s suspected exposure history, according to the available case contact tracing report (see Methods and Supplementary Methods section 1.1 for details in data processing). There were 126 infectors and 344 infectees with available information on complete exposure window as well as symptom onset times. Fig. 1 presented the epi-curves for the number of infectors and infectees identified over time based on their onset dates. We found 7-day moving window could ensure sufficient sample size for the temporal analysis on the estimation of these epidemiological parameters under forward and backward schemes, while the first and last moving windows were widened to capture the sporadic cases in the early and declining phases of the epidemic (Supplementary Tables 1-2). The details of the number of infectors and infectees that have complete exposure information, and number of transmission pairs in each time window of the study period can be found in Supplementary Methods section 1.2.

**Fig. 1.**
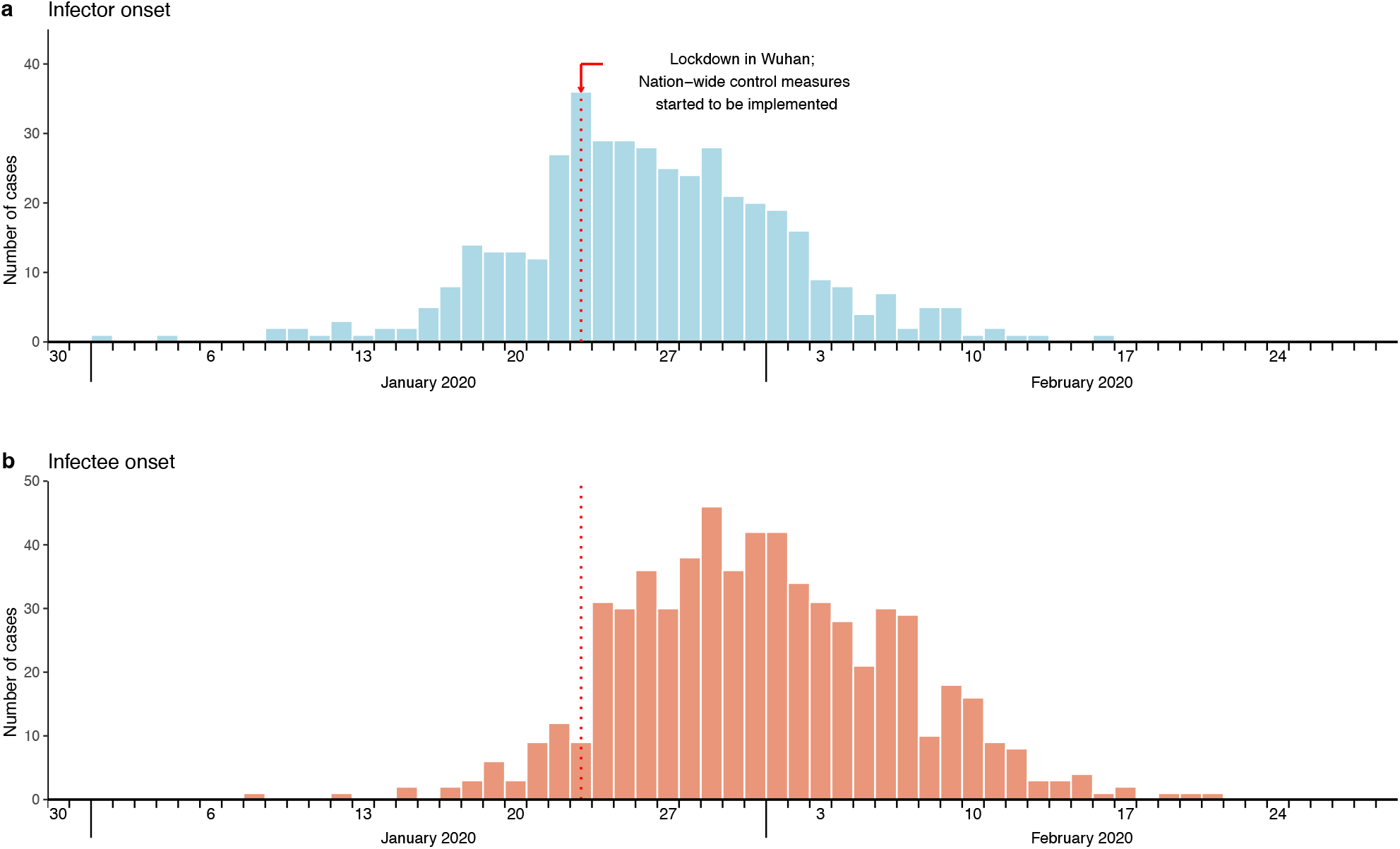
Infector-infectee specific symptom onset epi-curves from January 1 to February 29, 2020 in Mainland China. **a**, Epidemic curve based on symptom onset timing for the daily number of infectors. **b**, Epidemic curve based on symptom onset timing for the daily number of infectees.

### Temporal estimates of serial intervals, incubation periods and generation times

During the study period, the empirical forward SI decreased from a mean of 8.90 (interquartile range (IQR): 5.00 – 11.25) days to 2.68 (IQR: 0.00 – 6.00) days (Fig. 2a). The estimated mean backward IP of infectors increased from 3.77 (95%CI: 3.09, 4.53) days to 9.61 (8.14, 11.13) days (Fig. 2b), and the mean forward IP of infectees also increased from 5.39 (4.50, 6.30) days to 7.21 (6.36, 8.10) days (Fig. 2b). The mean empirical backward SI showed an increasing trend over time (Supplementary Fig. 1a), as well as the backward IP of infectee, while IP of infector referenced by infectee onset was increasing during the early phase and later became stable till the end of the study period (Supplementary Fig. 1b).

**Fig. 2.**
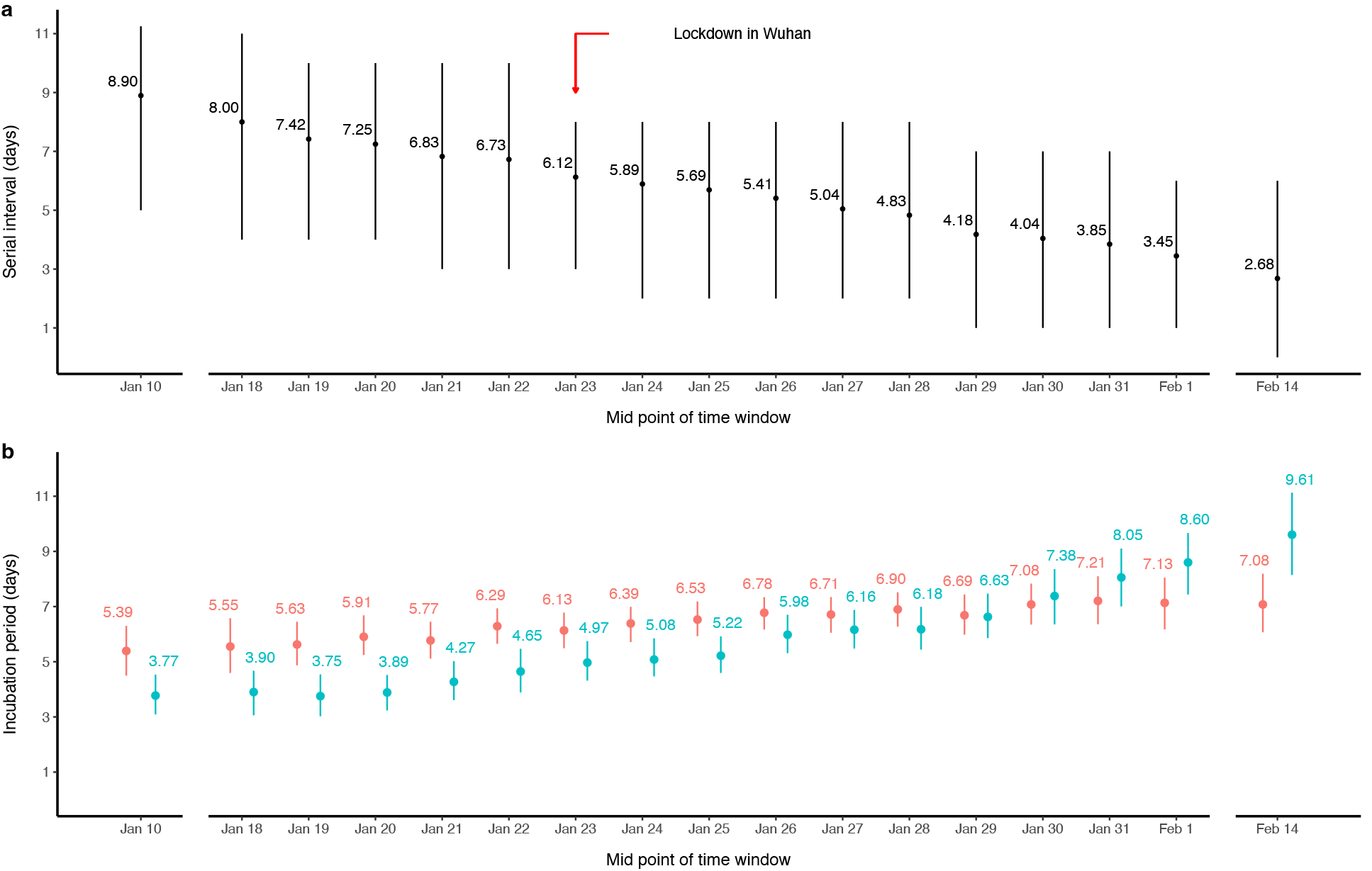
Temporal estimates of forward serial intervals (SIs) (a), forward incubation periods (IPs) of infectees, and backward IPs of infectors (b). **a**, Empirical mean and inter-quartile range (IQR) of forward SI in each moving window. The black dots and segments represent the empirical mean and IQR respectively. The red arrow indicates the timing of public health social measures (PHSMs) implemented since January 23, 2020. **b**, The estimated mean IP stratified by infector and infectee in each moving window. The dots and segments indicate the mean estimates and the corresponding 95% confidence intervals. The estimates for the forward IP of infectees and backward IP of infectors are presented in red and teal respectively.

The mean forward GT decreased from 7.27 (95%CI: 6.42, 8.07) to 4.21 (3.70, 4.74) days until January 29 and then increased slightly up to 5.20 (4.39, 6.02) days (Fig. 3a). While the estimated SD of forward GT decreased from 3.81 (2.84, 4.80) days on January 10 to 1.84 (1.38, 2.49) days on January 25 and then it increased to 3.65 (2.72, 4.51) days on February 14 (Fig. 3b). On the other hand, applying our estimation framework for backward GT, it was estimated that the mean backward GT ranged from 4.32 (3.87, 4.77) to 5.80 (5.25, 6.39) days (Supplementary Fig. 2).

**Fig. 3.**
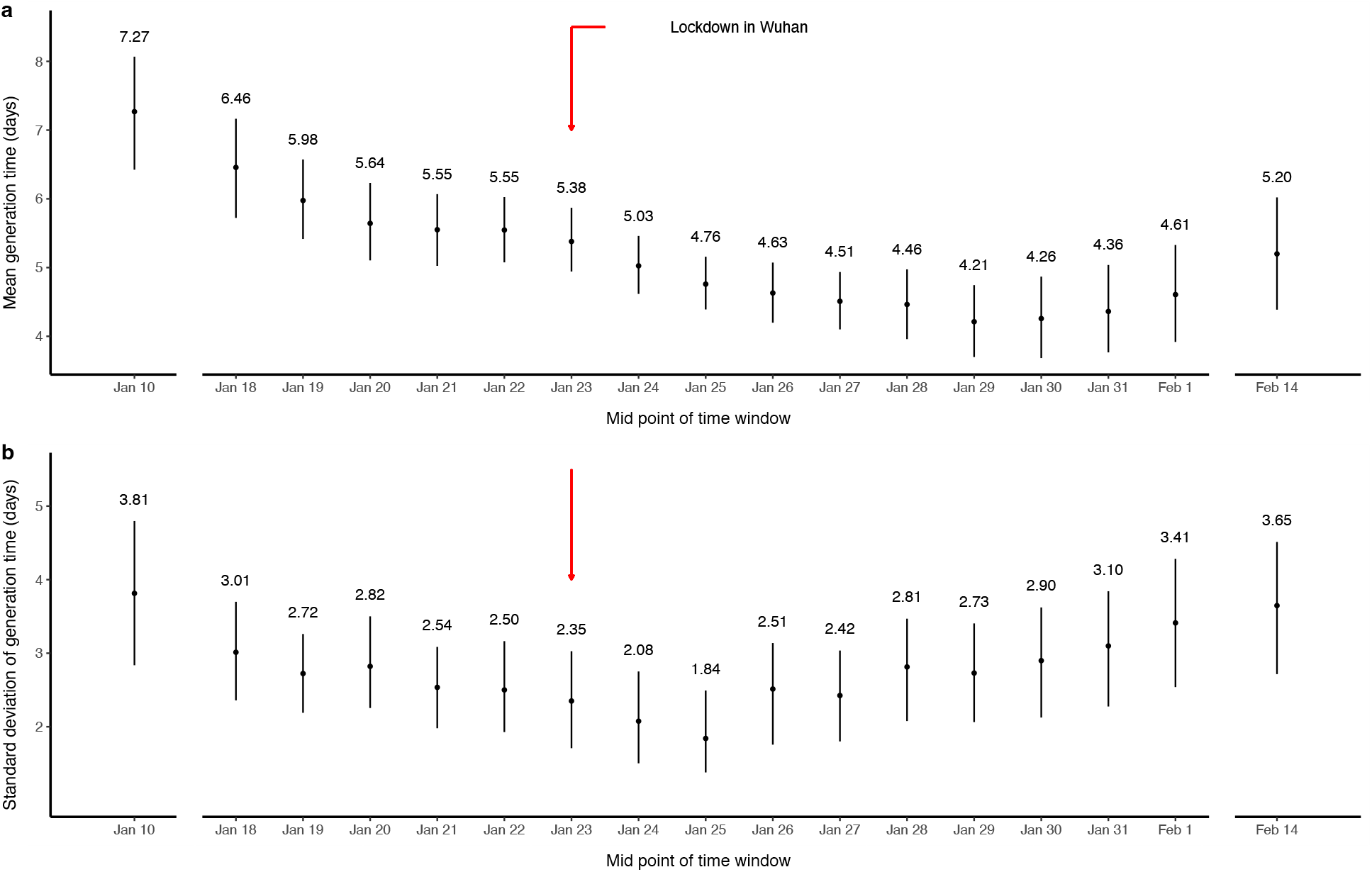
Temporal estimates of forward generation time (GT) distributions. **a**, The time-varying estimates of mean GT presented by the black dots with 95% confidence intervals (CIs) in vertical line-segments for each time window. **b**, The temporal estimates of standard deviation of GT presented by the black dots with 95% CIs in vertical line-segments for each time window. Red arrow indicates the implementation of public health social measures (PHSMs) since January 23, 2020.

### Sensitivity analysis and bias evaluation for generation time estimates

We compared the fittings by different choices of distributions for incubation periods and generation times respectively (Supplementary Table 3, Supplementary Fig. 3). The Weibull-distributed IPs of infectors and infectees and Log-normal distributed GT gave the lowest AIC values on the data for entire epidemics, while different choices of distributions for the forward GT showed similar AIC (difference < 5) in most of the moving windows. When the sampling biases in incubation period between infector and infectee at the temporal scale were not accounted, the estimated mean GT would be overestimated up to 17.83% in the early phase of the epidemic and underestimated up to 29.48% in the later phase with a decreasing pattern over the study period (Supplementary Fig. 4a). While estimated SD for GT would be overestimated up to 25.64% and underestimated up to 21.28% during the early and later phases respectively (Supplementary Fig. 4b).

We also compared the estimates under a model that considered the potential correlation between infector’s backward IP and forward GT (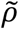), which suggested the correlations of 0.31(0.13 – 0.47) – 0.61(0.41 – 0.76) during the study period, as well as higher means (ranging from 5.12 (4.69 – 5.56) to 8.04 (7.25 – 8.89)) and higher standard deviations (ranging from 2.37 (1.89 – 2.87) to 4.41 (3.33 – 5.53)) of forward GT compared to the main result where independence between IP and GT was assumed (Supplementary Table 4). The changing patterns were consistent with main results (Supplementary Fig. 5). Similar estimates of GT were obtained when the correlation was assumed to be fixed at 0.25, 0.5 or 0.75 instead of being estimated by the model (Supplementary Table 4; Supplementary Fig. 5). However, our simulation study revealed that these estimates might suffer from bias (Supplementary Tables 9-11).

### Estimation of the basic and effective reproduction number

The basic reproduction number, *R*_0_, was estimated to be 1.95 (95% CI: 1.70, 2.26) given the exponential growth rate of 0.10 (0.08, 0.12), and forward GT distribution in the early part of the epidemic with a mean of 7.27 (6.42, 8.07) days and SD of 3.81 (2.84, 4.80) days. In contrast, when the backward GT distribution based on data from January 1 to 26, 2020 (the first moving window) was used instead, which had a mean of 4.93 (4.35, 5.53) days and SD of 2.99 (2.34, 3.57) days, *R*_0_ was estimated to be 1.58 (1.43, 1.74) which was underestimated by 18.97%.

The observed epi-curve of all cases onset showed the peak incidence was on January 29, 2020 (Fig. 4a). Based on this epi-curve, we estimated *R*_*t*_ by temporal GT distribution with reference to infector onset (red line in Fig. 4b) and effective SI distribution (Supplementary Fig. 6) with reference to infector onset (blue line in Fig. 4b) respectively. As shown in Fig. 4b, these two estimates and their corresponding confidence interval mostly overlap in the growing phase, and both declined to 1 at the end of January. But during the fading phase since February, the estimated *R*_*t*_ by temporal SI distribution was a little bit higher than the estimates by temporal GT distribution.

**Fig. 4.**
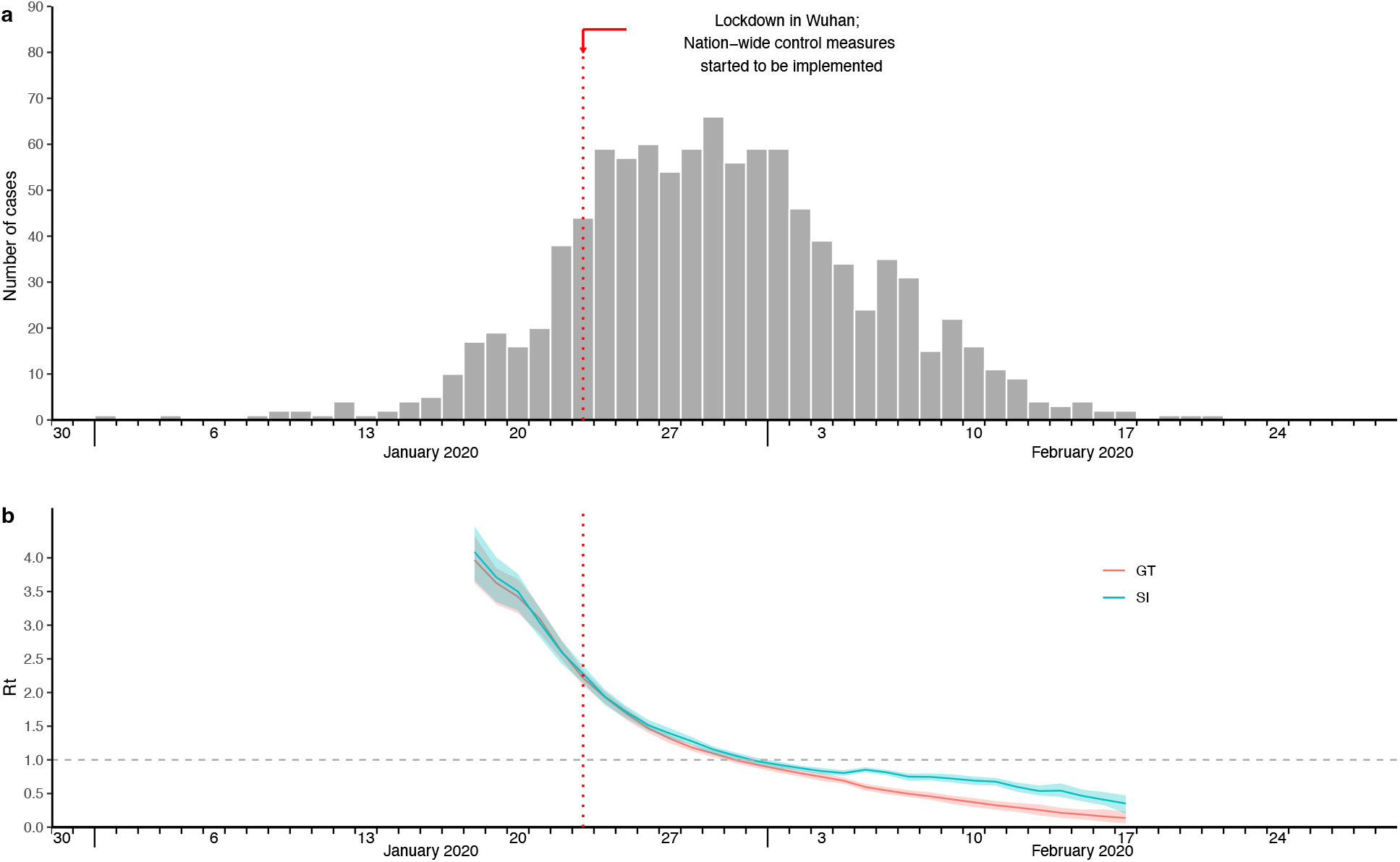
Epi-curve of observed onset times (a), and effective reproduction numbers estimated by temporal generation time (GT) and serial interval (SI) respectively (b). **a**, Epidemic curve of all cases symptom onset. **b**, Case-based effective reproduction numbers estimated based on epi-curve and temporal generation times (GT) with reference to infector onset, versus estimates based on epidemic curve and temporal serial intervals (SI) with reference to infector onset, shown as lines colored in red and teal respectively. Shaded areas correspond to 95% credible intervals of the estimates.

### Simulation results for inference of generation time

Park et al^11^ showed that the realized GT distribution over the simulated epidemics could be different from its intrinsic distribution, subject to sampling bias and susceptible dynamics in population. Based on our simulation study, our proposed inferential framework was able to recover the simulated values of realized GT, when the mean width of exposure window did not exceed the mean of intrinsic GT, and also below or approximately equal to the mean of intrinsic IP. Under such criteria, the proportions of 95% CI of estimated mean of realized GT covering simulated mean of realized GT ranged from 78% to 98% over all intrinsic GT setting (Supplementary Table 5), while the proportions of 95% CI of estimated SD of realized GT covering simulated SD of realized GT ranged from 80% to 100% based on 50 simulations (Supplementary Table 6), suggesting satisfactory recovery performance of our model. However, longer width of exposure window was associated with lower proportions of 95% CI of estimated value covering the simulated value, as well as larger bias especially overestimation in SD. When there were 1/3 of infector and infectees with completely missing exposure information, the proportions of 95% CI of estimated value of realized GT covering simulated value of realized GT would be generally lower, and bias in estimates were larger, compared to the situation when all infectors and infectees had complete exposure information (Supplementary Tables 7 – 8). Note the simulation of transmission data and estimation of GT were both under the assumption that IP and GT were independent. Besides, we further tested the reliability of using forward GT/SI to estimate effective reproduction number in the initial time window (*R*_*I*_) as a proxy of *R*_0_ (Supplementary Note section 2.2, Supplementary Table 9). We found that *R*_*I*_ would suffer from bias of 6% - 25% and -1% - 7% when forward SI distribution and forward GT distribution were used respectively, depending on the underlying intrinsic GT settings.

In another simulation study involving the intrinsic distribution of correlated forward IP and GT with a correlation coefficient of *ρ*, we tested the performance of our adjusted model that considered correlation between infector’s backward IP and forward GT (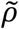) by estimating the realized correlation coefficient, the mean and SD of GT simultaneously (Supplementary Methods section 1.6). Simulation results suggested that the estimates were very sensitive to the width of exposure windows. The recovery performance was satisfactory when the mean width of exposure windows was 1 day (Supplementary Note section 2.3, Supplementary Tables 10-12), with the bias of <5% and proportion of 95% CI of estimate covering the realized value of >80% in almost all time windows especially when *ρ* ≤ 0.5. However, the exposure window with mean width of ≥ 4 days was associated with biased estimates (over-/under-estimation dependent on the parameters) (Supplementary Note section 2.3, Supplementary Tables 10 – 12). We thus reported the estimates under the assumed independence between IP and GT as the main result given the promising recovery performance in simulation studies.

## Discussion

We have obtained the time-varying estimates of generation times by incorporating the temporal changes in the estimates of serial intervals and incubation periods. Based on transmission pairs data, the mean generation time of COVID-19 was estimated to be around 7 days in the beginning of the epidemic in mainland China and the corresponding basic reproduction number was 1.95. In one month, the mean of generation time decreased to 4 – 5 days accounting for the effectiveness of public health and social measures (PHSMs) that were implemented to control transmission. Previous studies have estimated the mean generation time of COVID-19 in early 2020 to be 5.20 (95% CrI: 3.78, 6.78) days in Singapore^12^ and 5.70 days (95%CI: 4.80, 6.50) in mainland China^13^, which were both within the range of our temporal estimates in the growing-to-peak phase of the onset-based epi-curve. On the other hand, the mean of temporal GT was reduced to 4.21 (95% CI: 3.70, 4.74) days on January 29, which was consistent with the result reported by Li et al^14^ that the estimated mean GT decreased from 5.47 (95% CI: 4.57, 6.45) days in first generation to 4.25 (95% CI: 2.82, 6.23) days in successive generations with majority of the infectors exposed before and after January 23, 2020 respectively.

Depletion of susceptibles in the population due to high hazard of infection during the epidemic could temporally lead to reduction in mean of forward GT, which has been illustrated mathematically by Nishiura^15^ and further visualized by Champredon & Dushoff^16^ and Park et al^11^. However, an antibody seroprevalence study by Li et al^17^ estimated the weighted seroprevalence for Wuhan and provinces outside Hubei after the first wave in mainland China was only 4.43% (95% CI: 3.48%, 5.62%) and <0.1% respectively, indicating there should only be a limited degree of susceptible depletion that could lead to a reduction in the mean of the forward GT. It is more likely that the GT was shortened due to the implementation of nation-wide control measures on January 23, 2020^18^. Apart from lockdown in Wuhan, the nation-wide control measures included early detection and isolation of suspected cases, quarantine of close contacts, restricting opening time of public facilities and requiring mask wearing in public places^19^. Such control measures would reduce the forward infections from the infectors, hence shorten the mean GT, similar to the mean SI as illustrated in recent studies^11,18^. Besides, while the backward GT should have a consistently increasing pattern due to the nature of backward sampling^11,15,16^, the reduction in our backward estimated GT also suggested the impact of PHSMs on shortening GT (Supplementary Fig. 2).

We noted Sender et al^20^ investigated the unmitigated infectious profile during the early epidemic stage in mainland China based on 77 transmission pairs for which the infector developed symptoms before January 17, 2020. They estimated the mean GT of 9.7 (95%CI: 8.3, 11.2) days and SD of 6.9 (95%CI: 4.3, 10.1) days, with the estimated correlation coefficient between IP and GT of 0.75 (95%CI: 0.5, 0.9), and thus estimated *R*_0_ of 2.2 (95%CI: 1.9, 2.7). Our result considering correlated IP and GT meanwhile suggested a mean GT of 8.04 (95% CI: 7.25, 8.89) days, SD of GT of 4.41 (95% CI: 3.33, 5.53) days, and the estimated correlation coefficient of 0.41 (95%CI: -0.03, 0.64) considering the data before January 20, 2020. Despite different timeframe, while Sender et al adjusted for sampling bias with an assumed IP distribution and an assumed exponential growth rate of epidemic, we used the estimated forward/backward IP from our transmission pairs data, which might contribute to the difference in GT estimates and correlation estimates. Nevertheless, our result was generally comparable with that from Sender et al.

We have compared the effective reproduction number (*R*_*t*_) estimates by temporal GT distribution and SI distribution respectively, and showed that the estimates mostly overlap before the fading phase of the epidemic (Fig. 4b). During the fading phase, however, forward temporal SI would suffer from systematic bias of smaller mean and larger variance by overweighing the transmission pairs with shorter serial intervals ^11^, hence resulted in a higher *R*_*t*_ than that estimated by temporal forward GT. In particular, *R*_*t*_ here was limited to the epi-curve constructed from transmission pairs data instead that of all observed infections/ case-onsets in the first wave in mainland China. In fact, *R*_*t*_ in Fig. 4b was evaluated based on our observed data to compare the impact of time varying GT and SI under comparative settings, therefore initial *R*_*t*_ could not be directly compared with our estimated *R*_0_, which was calculated here based on the population-level growth rate using all case-onset data^21^. While in our simulation study (supplementary table 9) we tried using forward SI or GT distribution in the initial time window to obtain effective reproduction number as a proxy of *R*_0_, and found that using SI would suffer from substantially overestimation bias than using GT.

We conducted simulation studies to assess the performance of the proposed inferential framework by testing how efficiently the generation time could be recovered under known setting. For given mean generation time of 5-7 days^12–14^ and the 95% quantile of incubation period of 14 days for COVID-19, our model suggested promising estimates with >80% of 95% CIs (dependent on the parameters) covering the simulated values of realized GT when the intrinsic GT has a mean of 7 days and a SD of 4 days under the mean width of exposure windows of 7 days (14 days as maximum) (Supplementary Tables 5 – 6). However, our model might be sensitive to long exposure windows which resulted in poor recovery performance of forward generation time, especially when the exposure windows had a mean width larger than mean intrinsic IP (i.e., mean exposure width > 7 days while intrinsic mean IP of 6.5 days), or when the intrinsic generation time had a mean comparatively shorter than the mean width of exposure windows (Supplementary Tables 5 – 6). Infector/infectee with missing exposure windows would also have similar impact on GT estimates (Supplementary Tables 7 – 8). It is possible that the long width and the absence of information of exposure window led to more uncertainties in the estimates of incubation periods of infectors and infectees, and hence may lead to potential bias in the estimates of generation time.

One advantage of our method is that we allow time-varying estimations on epidemiological parameters, providing more information on transmission dynamics. The traditional approach usually estimates the generation time as a constant distribution over the whole epidemic, while our method can reflect the potential impact of PHSMs in reshaping the interval measures^18^. An additional advantage is that we have accounted for the sampling bias in each related interval parameter in the inferential framework. It is usually considered that the SI and GT share the same mean assuming the mean IP does not differ between infector and infectee. However, for the estimates at the temporal scale these assumptions were not often true, due to different sampling approach of infector and infectee along with the case characteristics. When the sampling bias in IP is not adjusted for, the mean GT will be overestimated and underestimated in the early and later phase of an epidemic respectively (Supplementary Fig. 4).

However, our study has some limitations. First, our analysis was limited to symptomatic cases, therefore the framework might not be directly adopted to the transmission pairs including asymptomatic infectors or infectees, and our results may be affected by selection bias as we only analysed a small proportion of all confirmed cases. Second, our method might be limited by the long width of exposure windows, which would lead to biased estimates of generation time especially when the intrinsic generation time is relatively short (Supplementary Tables 5 – 6). Based on our data, the average exposure widths for infector and infectee were 3.42 days and 5.87 days respectively, suggesting the possibility of biased estimates of forward GT as the estimated mean of GT was reduced to <5 days due to COVID-19 PHSMs. Third, we assumed incubation period and generation time were independent in our inferential procedure, which may not hold for example if there is an association between inoculum and incubation speed^22,23^, but pre-symptomatic transmission was observed^7,8^ and the literature does not have such clear evidence on the correlation between incubation period and generation time for COVID-19. Our method could be further extended to consider the correlation between incubation period and generation time (Supplementary Table 4), yet our simulation result suggested that those estimates might not be reliable since they were very sensitive to the width of exposure windows (Supplementary Tables 10-12). Fourth, the case definition might have changed during the study period. The diagnosis criteria and case definition in mainland China broadened over time^21^, therefore milder cases were more likely to be identified later in the epidemic. While previous studies indicated shorter time delay from infection to clinical outcome for severe COVID-19 cases^24^, and even for MERS and SARS^25,26^, this might lead to the increase in mean of the estimated forward incubation period. Moreover, our result was subject to recall bias which might affect the accuracy of the exposure information and onset timings in our data, hence the precision of our estimates.

In conclusion, we have developed a method to estimate forward temporal generation times of COVID-19 that accounts for the sampling bias and temporal variations in serial interval and incubation periods of infector and infectee, and provides improved and time-varying estimates. We identified potential biases in the estimates of generation times including sampling bias at temporal scale, emphasizing the importance of using more accurate GT estimation for understanding the time-varying transmissibility of COVID-19. The time-varying estimates of generation time could be crucial for better assessment of the disease dynamics and transmissibility, and could help to improve public health policies and mitigation strategies in real-time.

## Methods

### Data collection and characterizing epidemiological parameters

We used line list data reported by China’s municipal health commissions outside Hubei province from January 1 to February 29, 2020. The original data was extracted from the publicly available case reports provided by more than 200 municipal health commissions in Mainland China and reported in earlier studies^6,18,27^, and further integrated and compiled by Liu and colleagues^28^. The line-list data contains the information on the case demography (age, sex, occupation, residence place), exposure and contact history, onset and hospitalization dates, and potential transmission links in addition.

In this study, we reconstructed each possible transmission pairs by checking and compiling the information on epidemiology history, contact tracing reports and inter-relationship for these confirmed cases. We defined infectors as cases that had exposure history to risk areas or contagious person, and infected other cases within the same transmission chain/network, and the corresponding infectees as the cases who had contact history with the infector from his /her earliest exposure time until the isolation time. If infectees had more than one suspected infector, we considered the corresponding infector who contacted the infectee earlier during his/her infectious period; if more than one suspected infector contacted the infectee at the same day, we considered the corresponding infector, having closer and more frequent contacts with the infectee. For the cases in further complicated infection events with uncertain transmission paths, they were excluded from this study. We also investigated and constructed exposure windows for the cases with available exposure history and checked the symptom onset times as the time when the case developed symptoms or reported self-recognized discomfort for the first time during his/her illness history. See Supplementary Methods section 1.1 for details. Our study received ethical approval from the Institutional Review Board of the University of Hong Kong.

### Inferential framework of temporal generation times

The serial interval was found to be shortened over time by implementation of public health and social measures (PHSMs)^18,29^; further, forward and backward incubation period found to have different temporal patterns^11^. Therefore, the distribution of generation time based on the estimates of incubation periods and serial interval can vary over time. We considered the estimations under a 7-day moving window to ensure the sufficient sample size and to capture the temporal changes of these epidemiological parameters.

We first assessed different sampling approaches and identified the respective biases in these interval estimates. The backward sampling in estimating SI (i.e., referenced by infectee onset) would underestimate SI during the growth phase of the epidemic, because the transmission pairs with longer SI might be missed out as the corresponding infectees had not shown their illness onset yet. On the contrary, forward sampling in estimating SI (i.e., referenced by infector onset) would provide relatively reliable estimates, because the follow-up ended until every infectee onset was observed in that cohort of infectors. But the pairs with infector onset before the start of follow-up would be excluded by forward sampling scheme, which could lead to larger variance in the estimates accounting for very few observed pairs, especially during the growth phase. Therefore, the underlying problems brought by forward and backward sampling are in line with problems brought by left and right censoring (Figs. 5a – 5b). These issues also apply when estimating IP, where forward and backward sampling of IP is referenced by exposure time and onset time respectively. When a complete epidemic curve is observed, the retrospective backward and forward sampling of SI eventually result in same estimates as all cases are sampled. But at temporal scale (i.e., estimation with reference to a certain time period) the forward temporal SI would keep decreasing, whereas the backward temporal SI would keep increasing. Such change was attributed to the backward sampling bias in IP^11^, which suggested that the temporal SI may not be a good proxy of temporal GT.

**Fig. 5.**
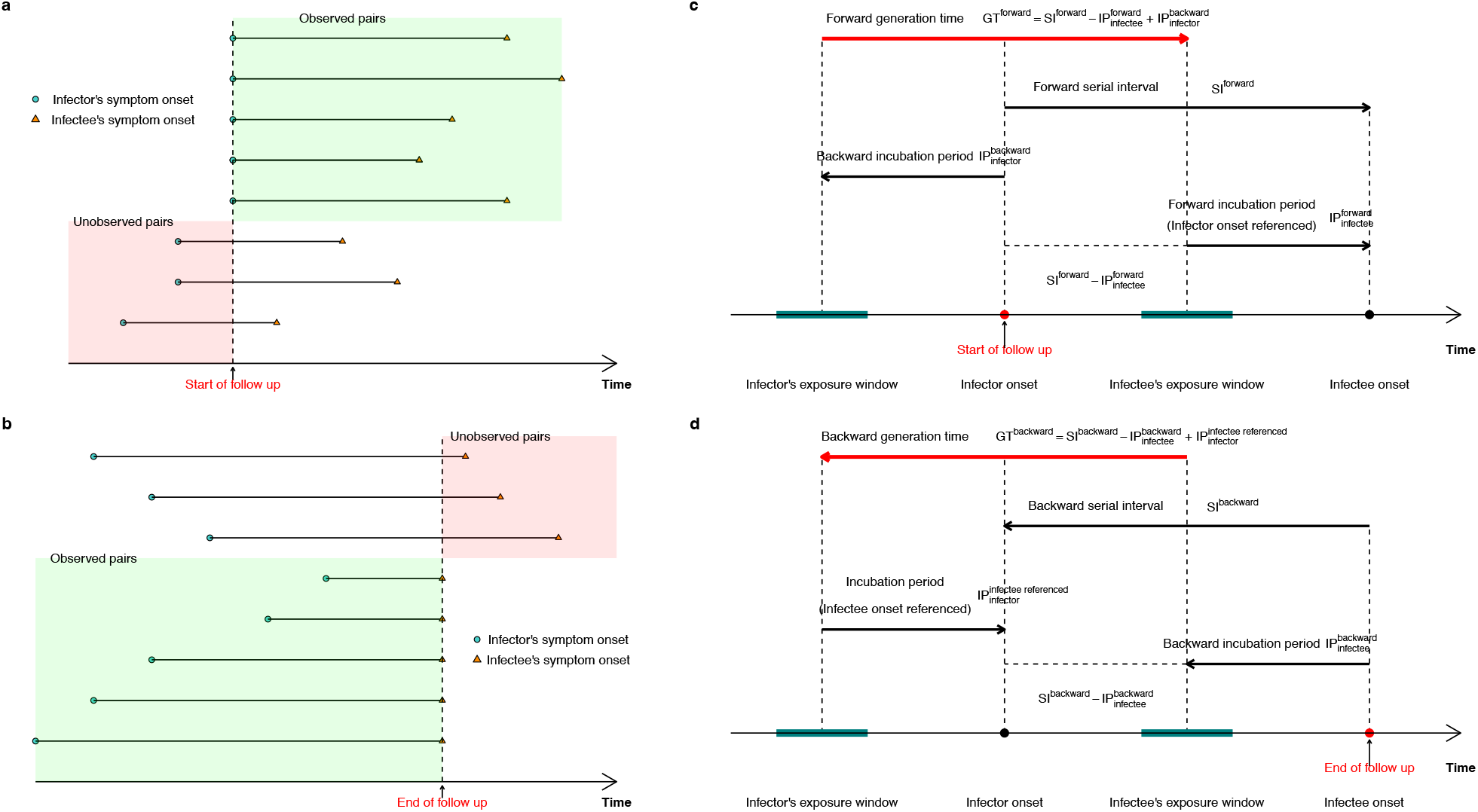
Censoring issues in sampling serial interval (SI) (a – b) and corresponding inferential frameworks for generation time (GT) (c – d). **a**, Forward sampling with reference point as the start of the event leads to left censoring issue. **b**, Backward sampling with reference point as the end of the event leads to right censoring issue. The biases are due to failure in observing the sample under these forward and backward schemes (as presented in the salmon colour shades). **c**, Inferential framework presented for forward GT. **d**, Inferential framework presented for backward GT. The inferential frameworks of GT have considered the inter-relationship among SIs and infector-infectee specific incubation periods (IPs).

In theory, GT should be referenced by infection times, which are rarely observed in practice. Consequently, we considered decomposing GT by respective forward and backward SIs as presented in Park et al^11^, and proposed the inferential frameworks for the estimates of forward and backward GTs based on the observations of these SIs and estimates of infector-infectee specific IPs as shown in Figs. 5c – 5d. For a given transmission pair *i*, the forward GT (*G*_*i*_) can be decomposed as the forward SI (*S*_*i*_) minus forward IP of infectee (*Y*_*i*_) plus backward IP of infector (*Z*_*i*_), i.e. *G*_*i*_ = *S*_*i*_ − *Y*_*i*_ + *Z*_*i*_. We assumed the IP of infectee is independent of the IP and GT of infector given the infection time of infectee, and the symptom onset time of infector is independent of infectiousness, thus the IP of infector is also independent of GT. Therefore, by assuming S, Y and Z are independently distributed, the probability density function of the GT for transmission pair *i* can be expressed as

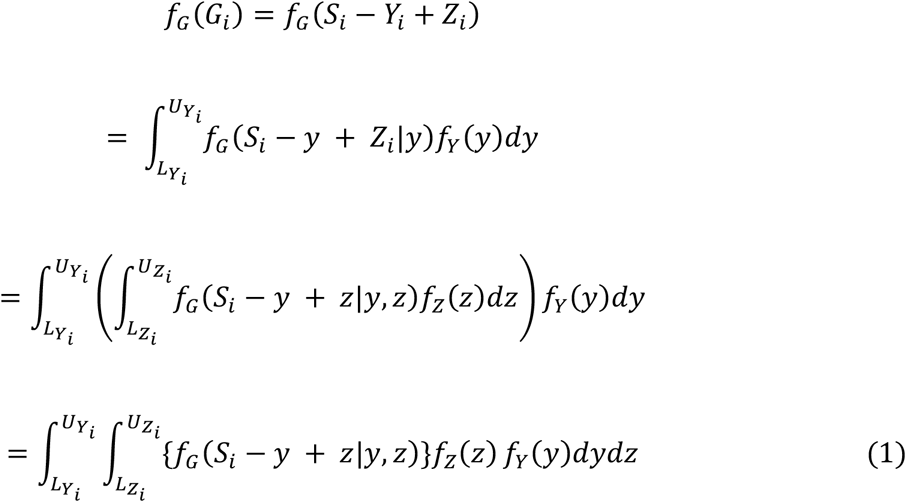

Where 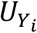 and 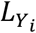 are the upper and lower bounds of IP for infectee, 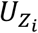 and 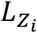 are the upper and lower bounds of IP for infector, *f*_*z*_(*z*) and *f*_*Y*_(*y*) are the probability density functions of infector’s backward IP distribution and infectee’s forward IP distribution respectively. Using Monte Carlo method, we can approximate this probability density function as

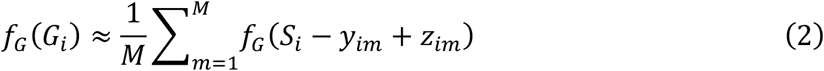

Where *M* is the number of Monte Carlo samples, *z*_*im*_ and *y*_*im*_ are the *m*-th Monte Carlo samples from *f*_*z*_(*z*) and *f*_*Y*_(*y*) for the *i*-th transmission pair respectively. Thus, given *N* transmission pairs, the likelihood function is given as

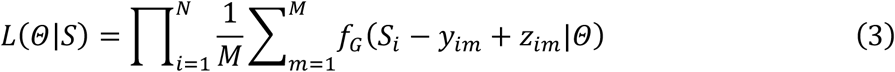

Where *Θ* is the parameter set of the GT and IP distributions. As the exact infection time is unobservable, we could infer the IPs from the exposure windows of the cases by fitting distributions on interval censored data, which could be used to generate Monte Carlo samples of IP and further evaluate the likelihood (Supplementary Methods section 1.3). On the other hand, when the dependence between IP and GT of infector was considered, we assumed the backward IP and forward GT of infector followed a bivariate normal distribution with a correlation coefficient 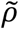 under logarithm scale. Similar approach considering the correlation between IP and GT was also used by Park et al^30^. The likelihood could be evaluated similarly by using the conditional distribution of forward GT given the Monte Carlo samples of backward IP of infector (Supplementary Methods section 1.6).

The 95% confidence interval (CI) was constructed by the percentile bootstrap method with 1000 bootstrapped samples. Statistical analyses were conducted using R version 4.0.4 (R Foundation for Statistical Computing). Visualization of estimations in inconsecutive time windows was implemented by R *ggbreak* package^31^.

### Sensitivity analysis on underlying distribution fitting

We first fitted three different distributions (Gamma, Log-Normal, Weibull) to infector’s and infectee’s incubation periods, and thus generated samples for GT which were further fitted by these three different distributions again. The results from the fitted distribution on GT samples with the lowest total Akaike Information Criterion (AIC) values over the moving windows were presented. We also evaluated the bias in GT estimates when the infectors and infectees were assumed to share the same IP distribution, where the sampling bias in infector and infectee’s IP were not adjusted for. We compared these estimates with main results that accounted for such sampling bias, and estimated the corresponding degree of overestimation/underestimation in each time windows.

### Estimating the basic and effective reproduction number

We referred to the previous estimate of epidemic growth rate, reported by Tsang et al^21^ as 0.10 (95% CI: 0.08, 0.12) for mainland China excluding Hubei province before Jan 23, 2020, and estimated the basic reproduction number (*R*_0_) using the forward GT estimates in the first time-window in the study period, where such forward GT distribution was an approximation of intrinsic GT distribution^16^, hence the calculated *R*_0_ has a better reflection of the infection spread at the early phase of the epidemic. We estimated *R*_0_ using the Lotka-Euler equation^2^:

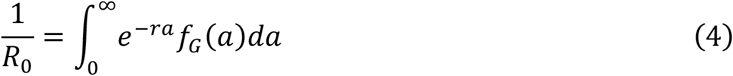

Where *r* is the growth rate, *f*_*G*_(*a*) is the generation time distribution. We simulated 1000 Monte Carlo samples of *r* and used our 1000 bootstrapped GT estimates to calculate *R*_0_. We use the term basic reproduction number to stress that over this period there were no population-wide interventions in place, and that all individuals were susceptible to infection.

Using the time-varying estimates of GT, we estimated the effective reproduction number *R*_*t*_, which shows the average number of secondary cases caused by one primary case at time *t*, accounting the population when some individuals may no longer be susceptible^32^. We used Wallinga & Teunis method^33^, a cohort based approach to estimate *R*_*t*_ via *EpiEstim* package in R (version 2.2-3)^34^. To compare the difference when using SI as a proxy of GT in evaluating transmissibility, we calculated *R*_*t*_ based on onset epi-curve and the time-varying estimates of GT and SI distributions respectively (Supplementary Methods section 1.4).

### Model validation by simulation studies

We have conducted several simulation studies to validate our proposed method. We first built an individual-based stochastic susceptible-infected-recovered (SIR) model with population size of 1000, 10 initial infected people, *R*_0_ equals to 2.5^11^. Given a Gamma-distributed intrinsic IP with a mean of 6.50 days and standard deviation (SD) of 3.50 days, we assessed the model performance under different distributions of intrinsic GT, where the intrinsic distribution indicates the original distribution at the initial phase of the epidemic^16^. During the progress of the epidemic, the distribution of realized GT may change due to high hazard rate of infection, particularly during the peak activity^15,35^. We assessed how our proposed framework could recapture the changes in mean and variance of the temporal realized GT during the epidemic progress. We tested three intrinsic GT settings of short (mean of 4 days, SD of 2 days), medium (mean of 7 days, SD of 4 days), and long (mean of 10 days, SD of 6 days) GT.

Besides, we also assessed how the width of exposure windows would influence the estimation accuracy. We assumed the width of exposure windows was uniformly distributed, and tested the recovery performance of parameters when the mean width of exposure windows was shorter than, equal to, and longer than the expected intrinsic GT (Supplementary Methods section 1.5). Furthermore, we assessed the recovery performance when 1/3 of infector and infectees did not have exposure information available (i.e. both earliest and latest exposure time were unknown) and further allowing no more than 1/3 of them partly missed exposure information (i.e. earliest exposure time unknown), as observed in our data, therefore in each simulation around 1/3 to 2/3 infector and infectees had complete exposure information in the medium GT setting.

On the other hand, we adopted the similar setting for the simulation studies for assessing the model performance which considered correlation between infector’s backward IP and forward GT. Focusing on the medium GT setting, we used the Log-Normal-distributed intrinsic IP with a mean of 6.50 days and standard deviation (SD) of 3.50 days, and Log-Normal-distributed intrinsic GT with a mean of 7 days and SD of 4 days during simulation, where they were correlated with a correlation coefficient *ρ* under the logarithm scale (Supplementary Note section 2.2). We tested the model performance under different *ρ* ∈ {0, 0.25, 0.50, 0.75} and mean width of exposure windows of 1, 4, 7 days.

## Data Availability

All the data used in the analysis will be available at Github (on acceptance):
https://github.com/DxChen0126/

https://github.com/DxChen0126/

## Data Availability

https://github.com/DxChen0126/

## Data availability

All the data used in the analysis will be available at Github (on acceptance): https://github.com/DxChen0126/

## Code availability

Statistical analyses were conducted using R version 4.0.5 (R Foundation for Statistical Computing, Vienna, Austria). Code will be available at Github (on acceptance): https://github.com/DxChen0126/

## Acknowledgements

The authors thank Julie Au for technical assistance. This project was supported by the Health and Medical Research Fund (project no. 20190712); a commissioned grant from the Health and Medical Research Fund from the Government of the Hong Kong Special Administrative Region; The Collaborative Research Scheme (Project No. C7123-20G) of the Research Grants Council of the Hong Kong SAR Government; and AIR@InnoHK administered by Innovation and Technology Commission. The funding bodies had no role in study design, data collection and analysis, preparation of the manuscript, or the decision to publish.

## Author contributions

All authors meet the ICMJE criteria for authorship. B.J.C., S.T.A. and D.C. conceived the study. D.C. and X.K.X. prepared the data. D.C., Y.C L. and S.T.A. developed the model. D.C. and Y.C L. conducted the data analyses. S.T.A., X.K.X., T.K.T., Z.D., L.W., P.W., E.H.Y.L., J.W. and B.J.C. interpreted the results. D.C., Y.C L. and S.T.A. wrote the first draft of the paper. B.J.C. and S.T.A. supervised the study. B.J.C. and S.T.A. acquired funding for the study. All authors provided critical review and revision of the text and approved the final version.

