## Supplementary Information for "Inferring time-varying generation time, serial interval and incubation period distributions for COVID-19"

### for 'Inferring forward generation time concerning temporal changes in incubation periods and serial intervals for COVID-19'

### 1. SUPPLEMENTARY METHODS

#### 1.1 Data Processing

The transmission pair data in this study was originated from COVID-19 case information reported by 27 provincial and 264 urban health commissions in mainland China during January 1, 2020 to February 29, 2020, outside Hubei province. Liu et al[1] had collected the original line list data and made it publicly accessible, including patient's encoding ID, age, sex, symptom onset date, quarantine date, hospitalization date, travel history, contact history and original information link (either from health commission website or local media website) that further contains occupation and residence place of the case if available. Ali et al[2] had first made use of mainland China's line list data and distinguished 677 infector-infectee transmission pairs to estimate the serial interval. In this study, we first referred to the dataset published by Ali et al for obtaining the transmission pair information, the further checked with the dataset published by Liu et al to examine the transmission link and obtain exposure information of the infector and infectee.

Our checking criteria for obtaining exposure information identifying infector and infectee is as follows:

1. The exposure history of a case is determined as

    i) If this case had been to risk areas (i.e. Wuhan/Hubei, places where COVID-19 cases had been reported), the first and last day in risk areas were regarded as the earliest and latest exposure time respectively;

ii) If this case did not have travel history but had contact history to Wuhan/Hubei personnel or other confirmed COVID-19 cases (who had earlier exposure history), the first and last day of contacting the suspected infector were regarded as the earliest and latest exposure time respectively;

iii) For family clusters, if index case's suspected infection source was travel history to Wuhan/Hubei, the infectee's earliest exposure time was the time when the index case returned to home; if index case's suspected infection source was contact history to contagious people, the infectee's earliest exposure time was the first time when the index case returned to home after contacting an contagious person (usually the same day); if index case were quarantined before infectee showing symptom onset, the infectee's latest exposure time was the time when index case started quarantine; if infectee developed symptoms before index case being quarantined, the infectee's latest exposure time was the same as symptom onset time;

iv) If a case had both been to risk areas and contacted to contagious people, the first day in risk areas and last day of contacting the contagious person were regarded as earliest and latest exposure time respectively.

2. A case is identified as infector if

i) He/she had travel history to risk areas, and he/she caused subsequent infections in his/her own network (i.e. other COVID-19 cases in his/her own network only had contact with this case as the suspected infection source);

ii) He/she had contact history to Wuhan/Hubei personnel, and he/she caused subsequent infections in his/her own network;

iii) He/she had contact history to other COVID-19 cases, and he/she caused subsequent infections in his/her own network.

3. A case is identified as infectee if he/she had contact history with another confirmed COVID-19 case, and did not have other suspected infection source.

4. We assumed a case's infectious period was within the time interval from a case's earliest exposure time to the quarantine time.

5. If a case had multiple suspected infectors, this case is regarded as the infectee connected to the infector that he/she first contacted within the infector's infectious period; if multiple suspected infectors were contacted at the same time, this infectee is connected to the infector that he/she had closer or more frequent contact with. (e.g. Let say case A contacted suspected infectors B and C at the same day. B was A's family member who lived with A, while C was A's friend. B would be considered as A's infector as A and B were expected to have more frequent contact compared to A and C.)

6. If a cluster (more than 2 people involved in a same contact network) had only one case with travel history to risk areas or contact history to contagious person from another contact network, then the rest of the cases in this cluster were all regarded as infectees of this case.

7. If a cluster had more than one case that could introduce infection to this cluster, then the transmission chain was unclear and the whole cluster was excluded from the study.

8. An infector does not necessarily have earlier onset date than infectee, as long as the infector had earlier infectious period than infectee's latest exposure time.

9. As we focus on one-to-one transmission pair, the infectee of a pair could be infector of another pair (i.e., if A infected B and B infected C, we will have two one-to-one transmission pairs in our data. For pair A-B, A is the infector while B is the infectee; for pair B-C, B is the infector while C is the infectee)

Based on the above criteria, we confirmed 629 one-to-one transmission pairs in this study.

#### 1.2 Determination of moving window and sample size

In our confirmed 629 one-to-one transmission pairs, there were 428 unique infectors showing symptom onset from January 1, 2020 to February 16, 2020. January 23 - 29, 2020 was the peak week of infector's symptom-onset-based incidence with 199 infectors onset, which accounted for more than one third of the total infectors; 107 and 122 infectors were onset before and after the peak week respectively. Thus, we defined the period of January 1-23, 2020 as the pre-peak phase of the epidemic, January 23 – 29 as the peak-timing phase of the epidemic, and January 29 – February 29, 2020 as the post-peak of the epidemic. On the other hand, the peak timing of infectees onset was about one week later than the infectors'. There were 231 infectees onset during January 29 - February 3, 2020, which accounted for more than one third of the total infectees. There were 213 and 185 infectees who had symptom onset before and after the corresponding peak week respectively.

There was a trade-off between sufficient sample size and timely reflection of the temporal changes in setting the length of time window for estimation. We set the window length of 7 days to ensure there were at least 20 infectors in each time window based on infector's onset, aiming to provide forward GT estimates with enough precision and sensitivity to temporal change. The sample size in each window were shown in Supplementary Table 1. On the other hand, we also conducted backward sampling based on infectee's onset to estimate backward GT. To ensure enough sample size in each time window, the backward moving window was different from forward moving window, as shown in Supplementary Table 2.

##### 1.3 Details of sampling procedure

We incorporated our inferential framework in a sampling procedure to obtain the GT samples as follows:

Step 1. Identify the transmission pairs whose infectors were onset in the given moving window. Suppose there are $N$ pairs provided observed serial interval values in the given time window $(S_1, S_2, \cdots, S_N)$.

Step 2. Among the identified transmission pairs, estimate the backward incubation period (IP) for infectors with complete exposure windows, and estimate the forward IP for infectees with complete exposure windows. Denote the estimated theoretical distributions for backward IP of infector and forward IP of infectee as $f_Z(z)$ and $f_Y(y)$ respectively.

Step 3. For the $i$-th transmission pair, do the following:

1) Revise or create exposure window for infector with complete or incomplete exposure window:
    a) The latest exposure time of infector is earliest time point among i) infector's latest exposure time (if available); ii) the earliest time point of all infectees' latest exposure time (if available); iii) the earliest onset time of infectees.

b) The earliest exposure time of infector is either i) infector's earliest exposure time (if available); ii) 21 days before infector's onset time; iii) 7 days before infector's latest exposure time when the latest exposure time exceed 21 days before onset time (to avoid extreme values).

2) Sample a random value $z_i$ for infector's IP from fitted distribution $f_Z(z)$ within the bound of infector's exposure window as determined above.

3) Revise or create exposure window for infectee with complete or incomplete exposure window:

   a) The earliest exposure time of infectee is latest time point among i) infectee's earliest exposure time (if available); ii) infector's possible infection time given $z_i$.

   b) The latest exposure time of infectee is the earliest time point among i) infectee's latest exposure time (if available); ii) infectee's onset time.

4) Sample a random value $y_i$ for infectee's incubation period from theoretical distribution $f_Y(y)$ within the bound of infectee's exposure window as determined above.

5) Based on $S_i$, $z_i$ and $y_i$, generate a forward GT sample as $G_i = S_i - y_i + z_i$.

Step 4. For the $i$-th transmission pair, generate $M$ (i.e. 1000) Monte Carlo samples for infector's IP (denoted as $z_{im}$) and infectee's IP (denoted as $y_{im}$) to obtain $M$ samples of forward GT. The probability density function of forward GT is approximated as $f_G(G_i) \approx \frac{1}{M}\sum_{m=1}^{M} f_G(S_i - y_{im} + z_{im})$, thus the likelihood over $N$ pairs can be expressed as: $L(\Theta) = \prod_{i=1}^{N} \frac{1}{M}\sum_{m=1}^{M} f_G(S_i - y_{mi} + z_{mi}|\Theta)$.

This sampling procedure can be incorporated with either forward or backward approach. Apart from main results based on forward approach, we also obtained backward empirical serial interval

(SI), forward IP of infector and backward IP of infectee as shown in Supplementary Figure 1. Based on these estimates, we implemented our sampling procedure to obtain the backward estimated GT as shown in Supplementary Figure 2. It was well noted that our estimated backward mean GT also decreased with the progress of the epidemic.

Further, under the forward approach scheme, we tested our model sensitivity by generating different GT samples through fitting different distributions on infector and infectee's IP in the sampling procedure, the goodness of fit of each estimation were summarized in Supplementary Table 3. And the fitted IP and GT mean and SD based on different distributions were shown in Supplementary Figure 3. Finally, we also evaluated the bias in estimated GT when the sampling bias between infector and infectee's IP was not acknowledged (i.e assuming infector and infectee share the same IP distribution), comparison with main result was shown in Supplementary Figure 4.

#### 1.4 Details of estimating effective reproduction number

The effective reproduction number $R_t$ estimated by Wallinga & Teunis method[3], which is also known as the case reproduction number. It focuses on a cohort of cases and calculates the likelihood of each case being infector that could have infected the rest of other cases in the given cohort. The calculation is given as follows:

$$p_{ij} = \frac{\omega(t_i - t_j)}{\sum_{k \neq i} \omega(t_i - t_k)} \qquad (1)$$

$$R_j = \sum_i p_{ij} \qquad (2)$$

Where $p_{ij}$ is the relative likelihood that case $i$ is infected by case $j$, which is the probability that case $i$ infected by case $j$ normalized by the sum of probabilities that case $i$ infected by cases other than case $i$. Note $\omega(\cdot)$ is the probability density function of the generation time distribution, and $t$ is the infection time. Then the effective reproduction number of case $j$ denoted as $R_j$, is the sum of all relative likelihoods that case $j$ being the infector of other cases.

We set the length of sliding window as 7 days to match with the moving windows for our temporal estimates. To ensure enough sample size for $R_t$ estimation in each moving window based on the onset epi-curve and forward GT estimates, our time windows started from the week of January 12 – 18 (sample size of 43 infection events), and ended on the week of February 11 – 17 (sample size of 35 infection events). We used *wallinga_teunis* function in R *EpiEstim* package (version 2.2-3)[4] to conduct the estimation.

The time-varying $R_t$ was not only based on the how the epidemic proceeded (i.e. epi-curve), but also the temporal GT. Thus, the $R_t$ in each time window was calculated based on the incidence as well as the corresponding temporal forward GT estimates. Note our first time window for forward GT estimation was January 1 – 20 and the last time window was January 30 – February 29, which were not with the length of 7 days. Therefore, the incidence during January 5 – 20 and the forward GT estimates based on the time window of January 1 – 20 would be used to estimate $R_t$ from January 18 to January 20. Similarly, the incidence during January 30 – February 14 and the forward GT estimates based on the time window of January 30 – February 29 would be used to estimate $R_e$ from February 5 to February 17.

For comparison, we followed the similar procedure to estimate $R_t$ based on onset-based epi-curve and the temporal forward SI, while we fitted the observed serial intervals by normal distribution to allow for negative values and used the fitted distribution as the proxy distribution for generation time in the equation for $p_{ij}$. The estimates of temporal SI are shown in Supplementary Figure 5.

#### 1.5 Details of stochastic SIR simulation setting

We referred to the earlier works[5,6] of Park and his colleagues to conduct the simulation study in this paper. The simulation was conducted by an individual-based stochastic susceptible-infectious-recovered (SIR) model assuming a fully connected population.

We simulated an epidemic started with 10 initial infectors in a fully connected network of 1000 people. Each initial infector would have infectious contacts with the network. The number of infectious contacts was assumed to be Poisson distributed with a mean of 2.5, where 2.5 was our assumed basic reproduction number in the entirely susceptible population in this simulation. The generation time for each infectious contact was assumed to follow a pre-specified Gamma distribution during simulation. If an infectee contacted with multiple infectors, only the infector who had earliest contact with the infectee would be considered to form the transmission pair with the infectee, and the corresponding contact time would be counted as the infectee's infection time.

The incubation period of infectees was assumed to follow Gamma distribution with a mean of 6.5 and SD of 3.5 days. The abovementioned distributions are so-call intrinsic distributions, while the distribution for infectious contact, generation time and incubation period would be regarded as realized distributions[5], which were subject to temporal changes due to stochasticity according to the SIR framework, for instance, the depletion in susceptible. The algorithm was updated by the order of infection time of each current infector, until no more infectious contact on a new infectee in the population was created, or the algorithm reached a predetermined time limit. Further details of the simulation algorithm can be found in Park et al 2020 & 2021[5,6].

Our model could recover the realized GT distribution theoretically. Since the realized GT distributions were unknown, we obtained the empirical mean and SD of realized GT over 1000 simulations and treated the average as the "true" mean and SD of realized GT. We assessed the model performance when intrinsic GT followed gamma distribution with a mean of 10 days and a SD of 6 days (long GT), with a mean of 7 days and a SD of 4 days (medium GT), and with a mean of 4 days and a SD of 2 days (short GT).

Besides, we also tested the impact of the width of exposure windows on the estimation accuracy. We simulated the lower bound $(L)$ and upper bound $(U)$ of the exposure window based on the infection time $(I)$ of the case, assuming $L$ and $U$ followed an Uniform distribution, i.e. $L = I - C_L$ and $U = I + C_U$, where $C_L$ and $C_U$ are uniformly distribution with the support of $(0, C)$, and thus the width of exposure window would have a mean of $C$ days. We would evaluate the estimation accuracy under the exposure windows with various width (i.e. $C \in \{1, 4, 7, 10\}$).

The recovery performance of model would be based on 50 simulated datasets. The estimates of parameters were obtained by maximum likelihood estimation.

##### 1.6 Sensitivity analysis on bivariate distribution of incubation period and generation time

In the main analysis the incubation period and generation time of infectors were assumed to be independent, which might not be realistic biologically. Thus, we further assess the estimates of forward GT distributions when we assumed the log-forward GT and log-backward IP of infector followed a bivariate normal distribution, hence the log-forward GT would follow a conditional normal distribution with a mean of $\mu_{G^*} + \frac{\sigma_{G^*}}{\sigma_{z^*}} \tilde{\rho}(z_{im}^* - \mu_{z^*})$ and a variance of $(1 - \tilde{\rho}^2)\sigma_{G^*}^2$ given the correlation coefficient $\tilde{\rho}$, the Monte Carlo sample of the log backward IP of infector $z_{im}^*$, and the parameters from the marginal distributions $\{\mu_{G^*}, \sigma_{G^*}, \mu_{z^*}, \sigma_{z^*}\}$. Where $G^*$ and $z^*$ denote the log value of GT and IP. Moreover, we evaluated the recovery performance of the model based on simulation studies considering different correlation coefficients between the intrinsic IP and GT (i.e. $\rho = \{0.00, 0.25, 0.50, 0.75\}$) and different width of exposure windows. Similar approach considering correlation between IP and GT was also used by Park et al[7].

### 2. SUPPLEMENTARY NOTE

#### 2.1 Simulation studies for testing performance of the inferential framework under independent incubation period and generation time

Supplementary Tables 4 – 5 showed the simulation results of applying our estimation method in obtaining mean and variance of forward GT, given different intrinsic GT settings and different length of exposure window, in which all cases had complete exposure information. Based on 50 simulations, we obtained satisfactory recovery performance of model when the mean width of exposure window was 7 days (i.e. maximum of 14 days) for medium and long GT setting, and when the mean width of exposure window was smaller than or approximately equal to the mean of intrinsic GT, as observed in short GT setting. When mean width of exposure window was 10 days for long GT setting, although it did not exceed mean intrinsic GT setting, recovery in SD had huge variations from 58% to 94%. We suspected that the long exposure window (e.g. mean width of 10 days) might not be informative enough to recover IP estimates especially when the intrinsic IP had a comparatively short mean (e.g. 6.5 days), which might affect the recovery performance in GT estimates as well. The above results suggested a condition for satisfactory recovery performance of GT, which was that the mean width of exposure window should be shorter than the minimum of the means of intrinsic IP or intrinsic GT. Here, we summarized the results of these simulation studies as presented in the Supplementary Tables 4 – 5.

Considering the situation where the mean width of exposure window was smaller than or equal to the mean of intrinsic GT, the proportions of 95% CI of estimated mean of realized GT covering "true" mean of realized GT ranged from 78% to 98% over all intrinsic GT setting, where the means of estimated means had bias of < 6.5%. In contrary, the estimated mean of realized GT had poor recovery of the "true" mean of realized GT indicated by the low proportions of 95% CI covering the "true" mean of realized GT (46% - 84%) and the large bias (-21.24% - 11.46%), especially when the mean width of exposure window was much longer than the mean of intrinsic GT.

On the other hand, the estimated SD of realized GT was more sensitive to the width of exposure windows. When the mean width of exposure window was smaller than or equal to the mean of intrinsic GT given the mean width of exposure window of ≤ 7 days, the proportions of 95% CI of estimated SD of realized GT covering "true" SD of realized GT ranged from 80% to 100% over all intrinsic GT setting, indicating satisfactory recovery of parameter. However, when the mean width of exposure window was larger than the mean of intrinsic GT, or when the mean width of exposure window was > 7 days, the proportions of 95% CI of estimated SD of realized GT covering "true" SD of realized GT ranged from 22% to 94% over all intrinsic GT setting. Moreover, longer exposure window was associated with overestimation of SD of realized GT. When the exposure windows had mean width of ≤ 4 days, the biases in estimated SD of realized GT were within ± 3%. However, overestimation of SD was obtained when the exposure windows had mean width of 7 days and when the mean width of exposure window was larger than the mean of intrinsic GT (with the bias of <10% generally). The biases in estimated SD of realized GT were >10% under the mean width of exposure window of 10 days.

We further assessed the recovery performance of parameters when 1/3 infector and infectees had completely missing exposure information under the medium instrinsic GT setting (Supplementary Tables 6 – 7), which was close to the situation observed in our data and our forward GT estimates. Similarly, shorter exposure window was associated with higher proportions of 95% CI of estimated values covering "true" value of realized GT, as well as smaller bias in magnitude. When the mean width of exposure windows were 7 days, the proportions of 95% CI of estimated mean of realized GT covering "true" mean of realized GT ranged from 72% to 84% over the moving windows, while that for SD ranged from 60% to 82%. The bias in estimated mean of realized GT was -8.11% - 3.75%, whereas the SD of realized GT was overestimated by >11%.

#### 2.2 Simulation studies for testing performance of using forward SI/GT to estimate initial effective reproduction number

We further assessed the recovery performance of basic reproduction number given the estimated forward SI/GT (GT was obtained with assuming independence between IP and GT). We used the effective reproduction number in the initial time window ($R_I$) as the proxy of basic reproduction number ($R_0$), where the empirical $R_I$ is calculated by multiplying $R_0$ and the proportion of susceptible in the population at the end time point of the initial time window, and we treated the mean of empirical $R_I$ over 1000 simulations as the reference $R_I$. On the other hand, the estimated forward GT and SI in the first time window were used to estimate $R_I$ by Wallinga & Teunis method[3] implemented in EpiEstim package[4], which would be compared with the reference $R_I$ based on 50 simulations. We also tested the recovery performance under different GT settings (long, medium, short) and different mean width of exposure windows.

We found forward SI would substantially provide overestimation of $R_I$ with bias ranging from 6% to 25% depending on the GT setting (supplementary table 9), along with poor recovery given the proportion of 95% CI of covering the reference $R_I$ of 68%, 32% and 6% under long, medium and short GT setting respectively. In contrast, using forward GT would lead to less bias in $R_I$ from -1% to 7% across different mean widths of exposure windows. The shorter width of exposure windows was associated with smaller bias in GT mean and SD, and hence a higher proportion of 95% CI of covering the reference $R_I$. Under the short GT setting, the proportion of 95% CI of covering the reference $R_I$ was 72% given the mean width of exposure windows of 1 day, but it dropped to <50% when mean width of exposure windows increased to >4 days. Yet, under the long and medium GT setting, the proportion of 95% CI of covering the reference $R_I$ would keep above 60%.

#### 2.3 Simulation studies for testing performance of the inferential framework under correlated incubation period and generation time

Focusing on the medium GT setting, we found that the recovery performance of the model considering correlated GT and IP was very sensitive to the exposure windows. Under the mean width of exposure windows of 1 day, the recovery performance was satisfactory especially when $\rho < 0.5$. In general, the estimated GT mean and GT standard deviation had biased of <5% (Supplementary Tables 10 - 11), whereas the bias in $\tilde{\rho}$ was more fluctuating despite that $\tilde{\rho}$ itself was in a small magnitude (Supplementary Table 12). The proportion of 95% CI of estimated GT mean covering the mean of realized GT could achieve >82% when $\rho \leq 0.5$, despite the proportion could possibly drop to 50% when $\rho = 0.75$ (Supplementary Table 10). The proportion of 95% CI of estimated GT standard deviation covering the "true" standard deviation of realized GT could achieve >88% when $\rho < 0.5$, and >72% when $\rho = 0.75$ (Supplementary Table 11). The proportion of 95% CI of estimated correlation covering the "true" realized $\tilde{\rho}$ was >78% (Supplementary Table 12).

However, when the exposure windows had a mean of >4 days, the recovery performance was inferior. The estimated GT mean suffered from overestimation especially under low $\rho$ and long exposure windows (Supplementary Table 10), where the bias could reach >5% and >10% under the mean width of exposure windows of 4 days and 7 days respectively when $\rho \leq 0.25$. In contrast, the estimated GT standard deviation suffered from underestimation under high $\rho$ and long exposure windows (Supplementary Table 11), specifically by >8% when $\rho \geq 0.25$ under the mean width of exposure windows of 4 days, and by >11% when $\rho \geq 0.5$ under the mean width of exposure windows of 7 days. Besides, the correlation could be severely underestimated under low $\rho$ and long exposure windows (i.e. >20% and >100% under the mean width of exposure windows of 4 days and 7 days respectively when $\rho = 0.25$) (Supplementary Table 12).

#### 2.4 Estimates of correlated generation time in transmission pairs in Mainland China

We estimated a higher GT mean and higher GT standard deviation based on the transmission pairs in Mainland China when correlation between forward GT and backward IP of infector was considered (Supplementary Fig. 5), and the estimated mean and standard deviation of GT were negatively associated under higher assumed $\tilde{\rho}$. Specifically, when $\tilde{\rho} = 0.25$, the estimated GT mean was longer by 0.71 – 1.44 days compared to the main result where correlation was not taken into account, while the estimated GT mean was longer by 0.06 – 0.94 days when $\tilde{\rho} = 0.75$ (Supp Table 4). Besides, the estimated GT standard deviation was longer by 0.18 – 0.46 days and 0.31 – 2.13 days compared to the main result when $\tilde{\rho} = 0.25$ and $\tilde{\rho} = 0.75$ respectively (Supplementary Table 4). On the other hand, we got similar estimates of GT when $\tilde{\rho}$ was estimated in the model instead, where the estimated $\tilde{\rho}$ ranged from 0.31 (0.13, 0.47) to 0.61 (0.41, 0.76) (Supplementary Table 4). Despite the poor recovery performance and biased estimates in simulation studies when correlation was considered, the trend of GT mean and standard deviation over the time windows were consistent with the trend in main result (Supplementary Fig. 5).

### 4. SUPPLEMENTARY FIGURES

**Supplementary Figure 1. Temporal empirical serial intervals (SIs) (a) and estimated incubation periods (IPs) referenced by onset of infectees (b). a,** Empirical mean and inter-quartile range (IQR) of backward SI in each moving window; the bold points represent the empirical mean, vertical line-segments are the IQR. **b,** The estimated mean IP stratified by infector and infectee in each moving window; the bold point are the mean estimates with 95% CI in vertical line-segments for forward IP of infectees (in red) and backward IP of infectors (in teal). Referencing to onset timing of infectees indicates the backward approach, study period was from January to February 2020 in mainland China.

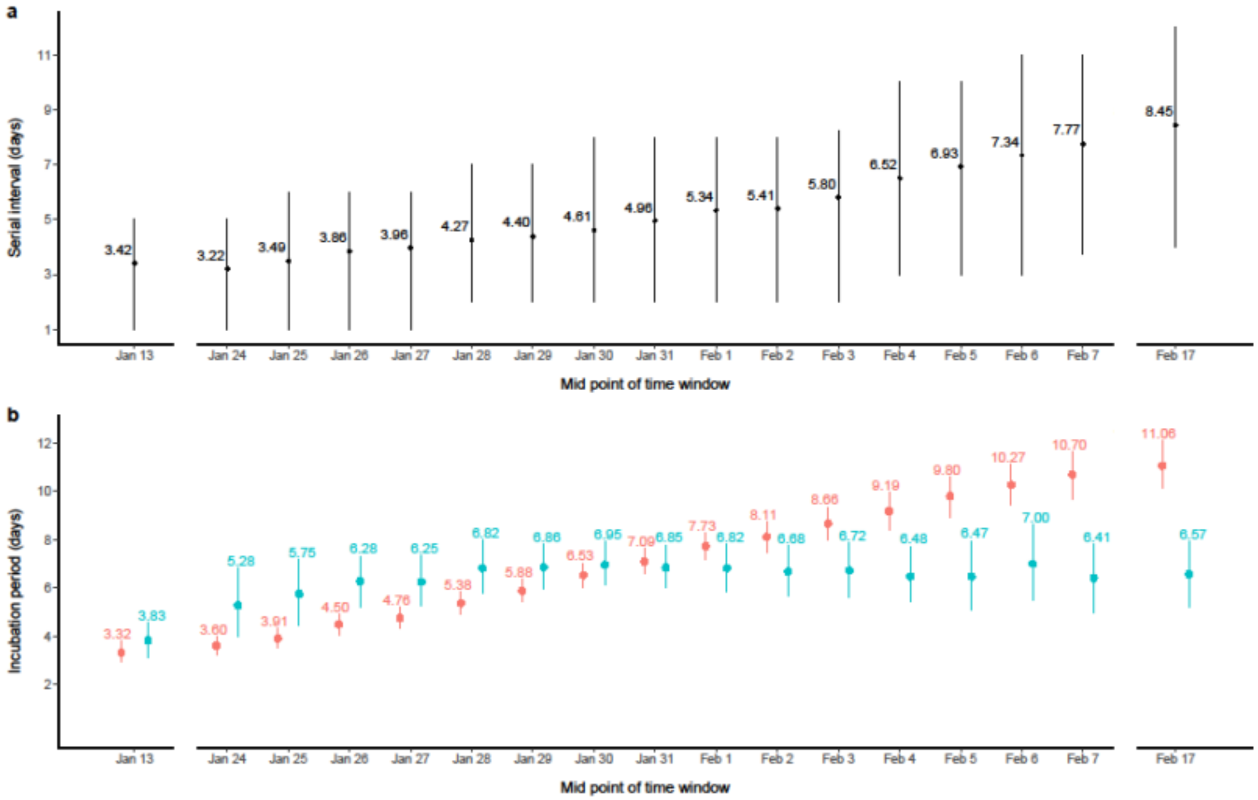

**Supplementary Figure 2. Temporal estimates of generation time (GT) distributions referenced by onset of infectees. a,** The time varying estimates of mean GT presented by the black dots with 95% CI in vertical line-segments for each time window. **b,** The temporal estimates of standard deviation of GT presented by the black dots with 95% CI in vertical line-segments for each time window. Referencing to onset timing of infectees indicates the backward approach, study period was from January to February 2020 in mainland China.

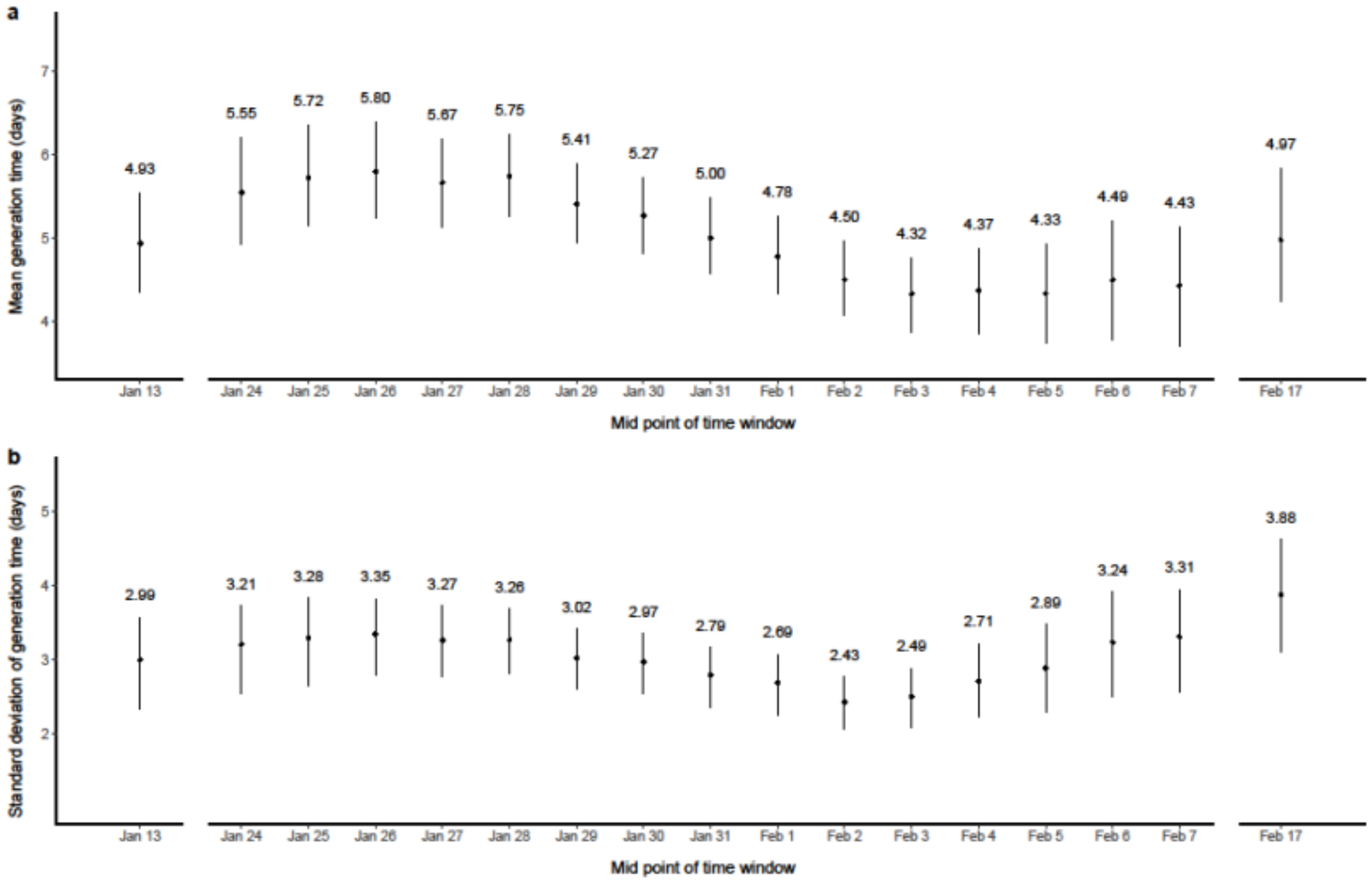

**Supplementary Figure 3. Fitting distributions on temporal infector-infectee stratified incubation periods (IPs) and generation time (GT) (forward approach).** **a,** Forward mean IP of infectee. **b,** Backward mean IP of infector. **c,** Forward mean GT. **d,** Standard deviation (SD) of forward IP of infectee. **e,** SD of backward IP of infector. **f,** SD of forward GT. The dots and lines showed the point estimates and the corresponding 95% confidence interval respectively, while the estimates fitted by Gamma, Log-Normal and Weibull distributions were shown in red, green and blue colours respectively. All estimations were with reference to onset timing of infectors, indicating forward approach.

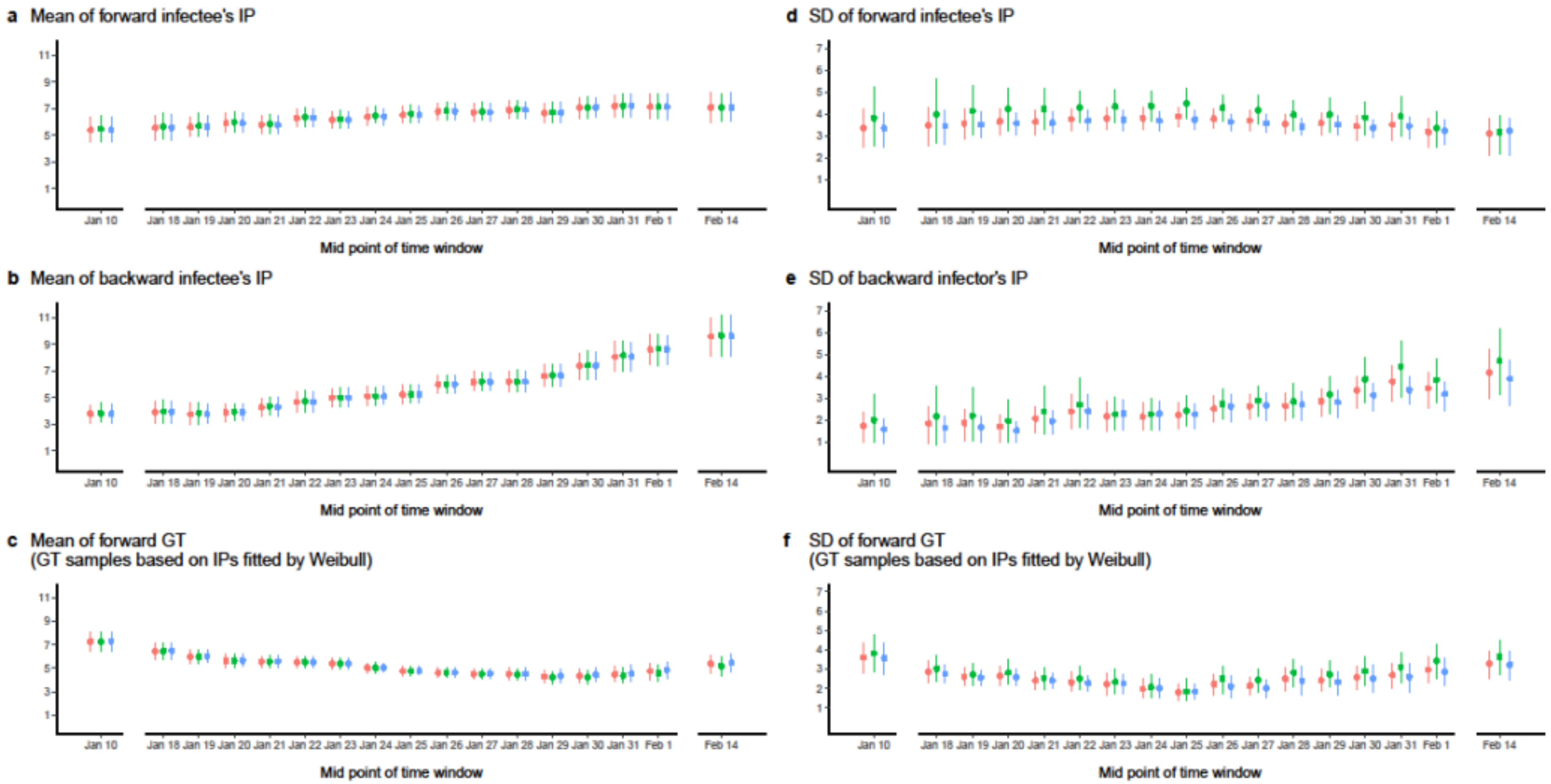

**Supplementary Figure 4. Comparison of generation time (GT) estimates with/without sampling bias in incubation period taken into account. a,** Comparisons in estimated mean forward GT. **b,** Comparisons in estimated standard deviation (SD) of forward GT. Dots showed point estimates, lines showed 95%CI of point estimates, colors red and teal represent GT was sampled based on estimated IP ignoring sampling bias between infector and infectee, and based on acknowledging the difference (main results) respectively.

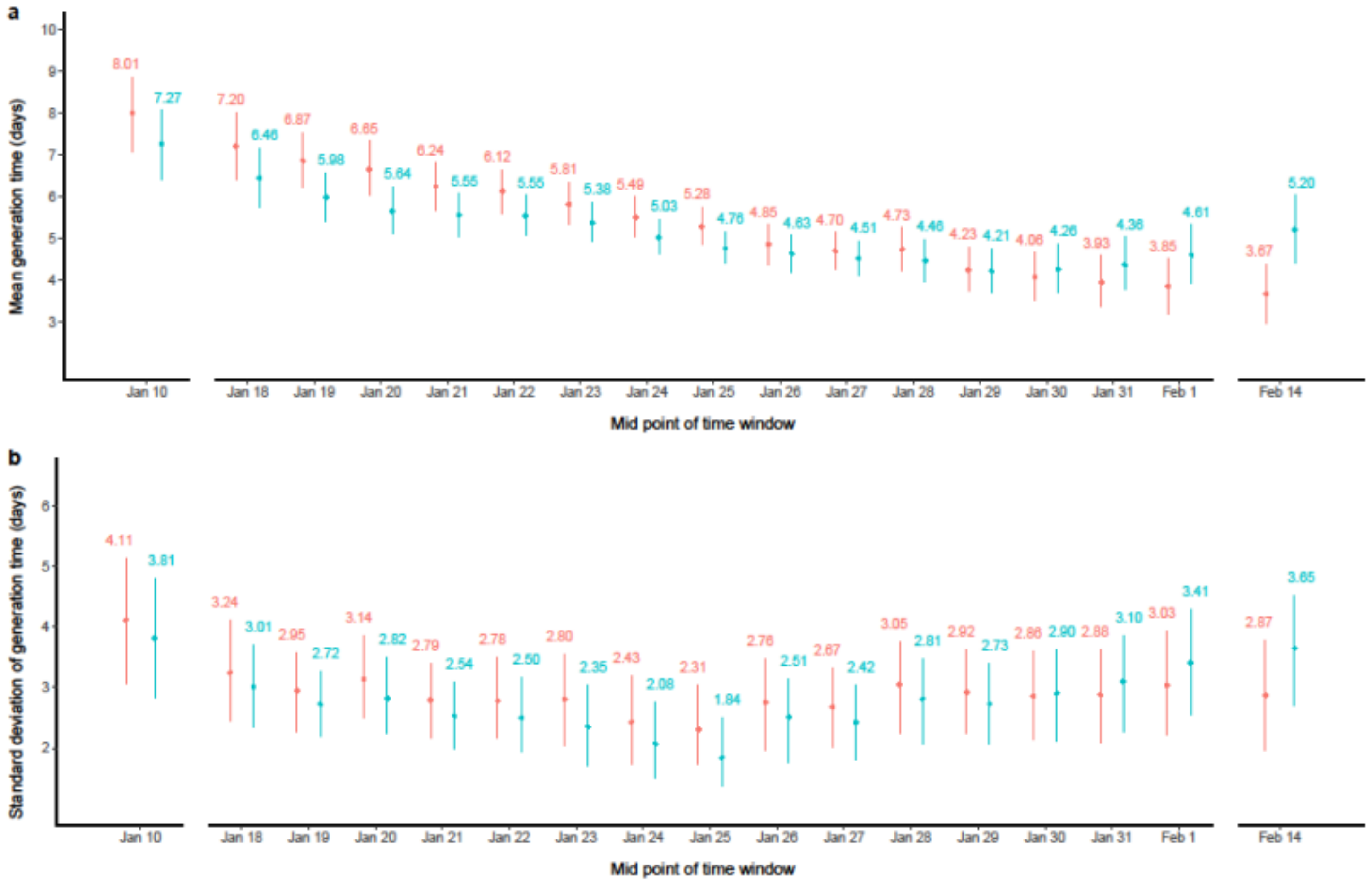

**Supplementary Figure 5. Comparison of generation time (GT) estimates with/without correlation between GT and incubation period (IP) taken into account. a,** Comparisons in estimated mean forward GT. **b,** Comparisons in estimated standard deviation (SD) of forward GT. Dots showed point estimates, lines showed 95%CI of point estimates, colors red, sand, green represent GT estimated based on fixed correlation coefficient $\rho$ at 0.25, 0.50, 0.75 respectively in the model; color in blue represent GT estimated based on the model that simultaneously estimate $\rho$ with mean and SD; color in purple represent GT estimates based on assumed independence of GT and IP in the model (main results).

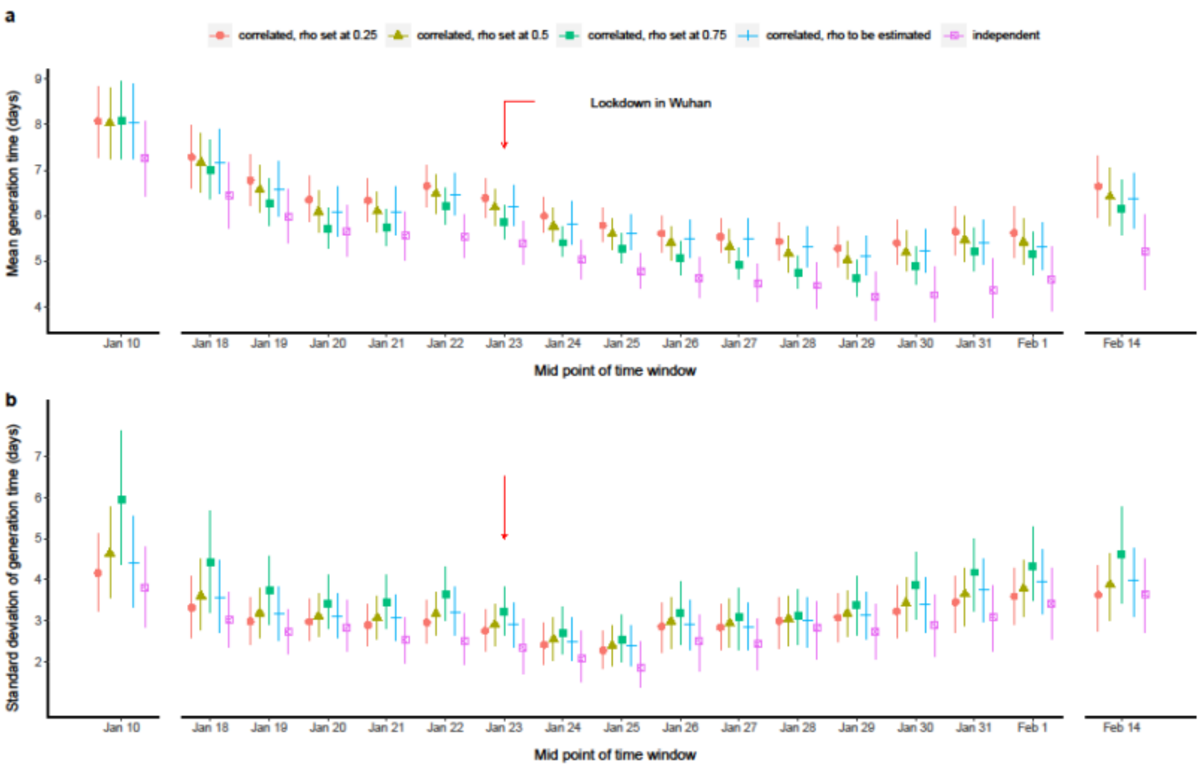

**Supplementary Figure 6. Temporal estimates of serial interval (SI) distributions referenced by onset of infectors. a,** The time varying estimates of mean SI presented by the black dots with 95% CI in vertical line-segments for each time window. **b,** The temporal estimates of standard deviation of SI presented by the black dots with 95% CI in vertical line-segments for each time window. The SI data were fitted by normal distribution. Referencing to onset timing of infectors indicates the forward approach, study period was from January to February 2020 in mainland China.

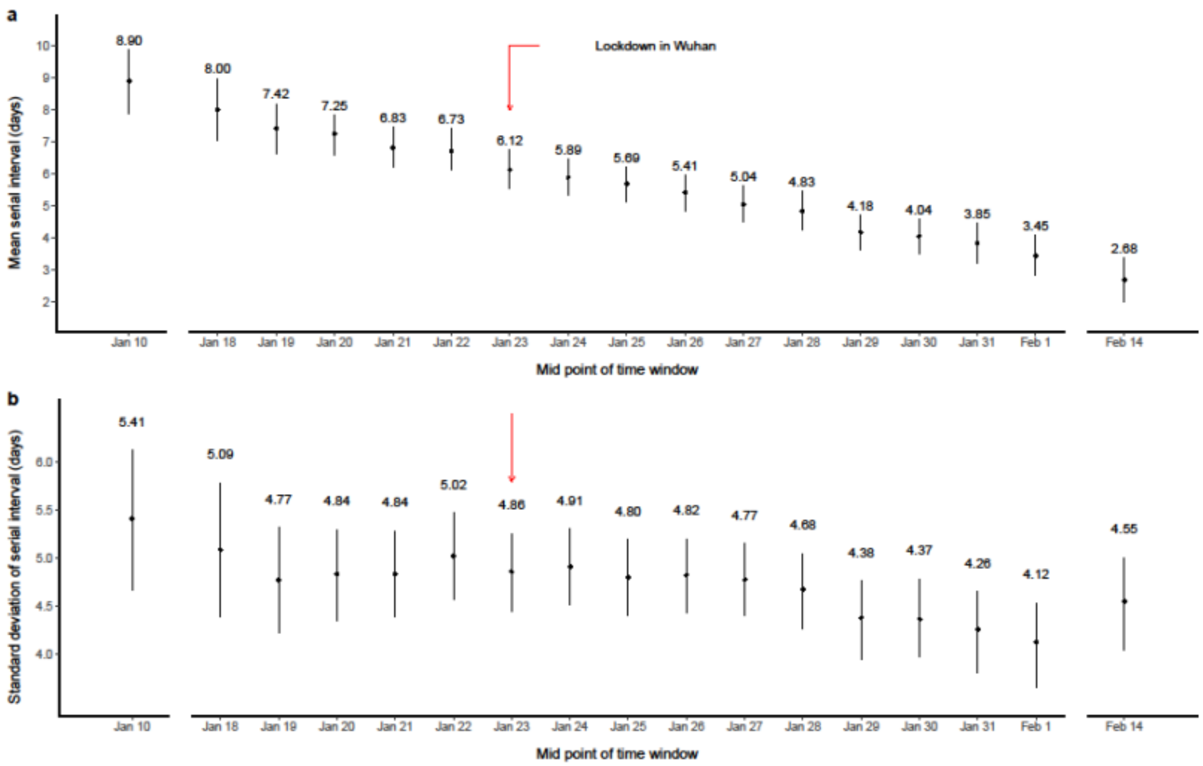

### 5. SUPPLEMENTARY TABLES

**Supplementary Table 1. Sample size in each time moving window for forward generation time (GT) estimation**

| Time window | All infectors with complete exposure | All infectees with complete exposure | One-to-one transmission pairs identified |
|---|---|---|---|
| **Jan 01 - Jan 20** | 25 | 62 | 116 |
| **Jan 15 - Jan 21** | 21 | 62 | 110 |
| **Jan 16 - Jan 22** | 24 | 90 | 146 |
| **Jan 17 - Jan 23** | 32 | 118 | 191 |
| **Jan 18 - Jan 24** | 36 | 134 | 218 |
| **Jan 19 - Jan 25** | 41 | 147 | 248 |
| **Jan 20 - Jan 26** | 44 | 155 | 257 |
| **Jan 21 - Jan 27** | 47 | 167 | 275 |
| **Jan 22 - Jan 28** | 51 | 173 | 291 |
| **Jan 23 - Jan 29** | 61 | 176 | 295 |
| **Jan 24 - Jan 30** | 59 | 157 | 270 |
| **Jan 25 - Jan 31** | 52 | 145 | 265 |
| **Jan 26 - Feb 01** | 50 | 123 | 231 |
| **Jan 27 - Feb 02** | 46 | 110 | 218 |
| **Jan 28 - Feb 03** | 43 | 87 | 188 |
| **Jan 29 - Feb 04** | 37 | 76 | 166 |
| **Jan 30 - Feb 29** | 34 | 70 | 163 |

**Supplementary Table 2. Sample size in each time moving window for backward generation time (GT) estimation**

| Time window | All infectors with complete exposure | All infectees with complete exposure | One-to-one transmission pairs identified |
|---|---|---|---|
| **Jan 01 - Jan 26** | 27 | 87 | 145 |
| **Jan 21 - Jan 27** | 28 | 97 | 157 |
| **Jan 22 - Jan 28** | 34 | 110 | 186 |
| **Jan 23 - Jan 29** | 40 | 131 | 220 |
| **Jan 24 - Jan 30** | 46 | 147 | 247 |
| **Jan 25 - Jan 31** | 43 | 149 | 258 |
| **Jan 26 - Feb 01** | 50 | 157 | 270 |
| **Jan 27 - Feb 02** | 52 | 160 | 268 |
| **Jan 28 - Feb 03** | 48 | 157 | 269 |
| **Jan 29 - Feb 04** | 50 | 153 | 259 |
| **Jan 30 - Feb 05** | 45 | 135 | 234 |
| **Jan 31 - Feb 06** | 47 | 126 | 228 |
| **Feb 01 - Feb 07** | 45 | 112 | 215 |
| **Feb 02 - Feb 08** | 36 | 93 | 183 |
| **Feb 03 - Feb 09** | 33 | 78 | 167 |
| **Feb 04 - Feb 10** | 34 | 70 | 152 |
| **Feb 05 - Feb 29** | 34 | 66 | 157 |

**Supplementary Table 3. AIC values of estimated forward incubation periods (IPs) of infectee, backward IPs of infector, and forward generation time (GT) with different assumed distributions.** AIC values were compared between estimations fitted by Gamma, Weibull and Log-Normal distributions by time windows. The AIC of forward GT under different distributions were based on best fitted Weibull distributed IPs. The lowest AIC values in each time window were marked in bold.

| | Forward IP of infectee | | | Backward IP of infector | | | Forward GT | | |
|---|---|---|---|---|---|---|---|---|---|
| **Time window** | **Gamma** | **Weibull** | **Log-Normal** | **Gamma** | **Weibull** | **Log-Normal** | **Gamma** | **Weibull** | **Log-Normal** |
| **Jan 01 - Jan 20** | **184.35** | 184.49 | 186.72 | 67.62 | **65.82** | 70.38 | 648.28 | 650.91 | **648.16** |
| **Jan 15 - Jan 21** | **187.85** | 187.98 | 190.24 | 59.79 | **57.62** | 62.75 | **583.26** | 583.27 | 584.60 |
| **Jan 16 - Jan 22** | **270.98** | 271.18 | 274.38 | 67.20 | **65.00** | 70.26 | **751.52** | 752.36 | 752.60 |
| **Jan 17 - Jan 23** | **356.32** | 356.48 | 360.72 | 81.17 | **77.98** | 85.14 | 982.44 | 986.12 | **981.62** |
| **Jan 18 - Jan 24** | **395.21** | 395.55 | 399.55 | 108.84 | **106.88** | 112.59 | 1118.99 | 1122.89 | **1118.32** |
| **Jan 19 - Jan 25** | 449.92 | **449.83** | 454.53 | **136.02** | 136.31 | 138.82 | 1281.56 | 1286.49 | **1279.32** |
| **Jan 20 - Jan 26** | **447.96** | 448.08 | 452.14 | **137.69** | 140.37 | 137.69 | 1305.22 | 1312.48 | **1302.19** |
| **Jan 21 - Jan 27** | 471.22 | **470.03** | 477.77 | **138.71** | 141.28 | 139.02 | 1366.53 | 1373.67 | **1363.85** |
| **Jan 22 - Jan 28** | 498.69 | **496.99** | 506.26 | **156.67** | 158.03 | 158.44 | 1426.03 | 1431.09 | **1424.69** |
| **Jan 23 - Jan 29** | 523.78 | **521.53** | 531.45 | **204.72** | 208.07 | 205.78 | 1478.44 | 1482.87 | **1478.40** |
| **Jan 24 - Jan 30** | 451.48 | **449.67** | 458.07 | **209.94** | 211.57 | 212.04 | **1336.35** | 1337.61 | 1339.10 |
| **Jan 25 - Jan 31** | 402.07 | **399.67** | 408.61 | **182.93** | 185.26 | 183.91 | 1330.54 | 1338.15 | **1326.75** |
| **Jan 26 - Feb 01** | 311.40 | **311.18** | 314.96 | **186.67** | 186.85 | 189.05 | 1157.73 | 1162.52 | **1155.53** |
| **Jan 27 - Feb 02** | 282.59 | **281.91** | 286.91 | 182.80 | **180.40** | 186.84 | 1105.83 | 1111.01 | **1103.20** |
| **Jan 28 - Feb 03** | 231.27 | **230.94** | 234.55 | 182.73 | **179.83** | 187.05 | 970.73 | 974.84 | **968.56** |
| **Jan 29 - Feb 04** | **184.94** | 185.96 | 186.21 | 150.96 | **148.88** | 153.31 | 866.46 | 869.38 | **865.80** |
| **Jan 30 - Feb 29** | **102.34** | 103.05 | 103.00 | 134.14 | **132.38** | 136.76 | **886.33** | 888.80 | 886.49 |
| **Sum of AIC over all time window** | 5752.37 | **5744.52** | 5826.07 | 2388.60 | **2382.53** | 2429.83 | 18596.25 | 18664.47 | **18579.17** |

**Supplementary Table 4. Estimated mean and standard deviation of GT when incorporates correlation between infector IP and GT in the model.** CI: Confidence Interval; $\tilde{\rho}$: correlation coefficient between backward incubation period of infector and forward generation time.

| **Estimate, (95% CI)** | **$\tilde{\rho}$ to be estimated in the model** | | | **$\tilde{\rho}$ fixed at 0.25** | | **$\tilde{\rho}$ fixed at 0.50** | | **$\tilde{\rho}$ fixed at 0.75** | |
|---|---|---|---|---|---|---|---|---|---|
| **Time window** | **Mean** | **Standard deviation** | **$\tilde{\rho}$** | **Mean** | **Standard deviation** | **Mean** | **Standard deviation** | **Mean** | **Standard deviation** |
| **Jan 01 - Jan 20** | 8.04 (7.25, 8.89) | 4.41 (3.33, 5.53) | 0.41 (-0.03, 0.64) | 8.08 (7.28, 8.85) | 4.16 (3.23, 5.12) | 8.03 (7.25, 8.80) | 4.64 (3.56, 5.76) | 8.09 (7.25, 8.96) | 5.95 (4.37, 7.62) |
| **Jan 15 - Jan 21** | 7.17 (6.50, 7.90) | 3.57 (2.73, 4.47) | 0.48 (0.17, 0.68) | 7.28 (6.61, 8.00) | 3.31 (2.60, 4.09) | 7.16 (6.52, 7.82) | 3.60 (2.79, 4.50) | 7.01 (6.38, 7.67) | 4.40 (3.19, 5.67) |
| **Jan 16 - Jan 22** | 6.57 (6.00, 7.19) | 3.17 (2.51, 3.82) | 0.49 (0.30, 0.65) | 6.77 (6.23, 7.32) | 2.99 (2.42, 3.54) | 6.56 (6.07, 7.11) | 3.18 (2.58, 3.79) | 6.26 (5.80, 6.80) | 3.73 (2.90, 4.56) |
| **Jan 17 - Jan 23** | 6.07 (5.55, 6.64) | 3.10 (2.60, 3.67) | 0.51 (0.33, 0.66) | 6.35 (5.88, 6.87) | 2.97 (2.51, 3.53) | 6.08 (5.64, 6.54) | 3.10 (2.61, 3.66) | 5.70 (5.30, 6.16) | 3.42 (2.81, 4.10) |
| **Jan 18 - Jan 24** | 6.09 (5.58, 6.63) | 3.07 (2.52, 3.63) | 0.50 (0.32, 0.65) | 6.33 (5.86, 6.80) | 2.90 (2.39, 3.40) | 6.09 (5.65, 6.52) | 3.06 (2.54, 3.60) | 5.73 (5.34, 6.14) | 3.44 (2.80, 4.11) |
| **Jan 19 - Jan 25** | 6.45 (6.02, 6.93) | 3.19 (2.66, 3.80) | 0.52 (0.34, 0.67) | 6.65 (6.20, 7.11) | 2.96 (2.46, 3.48) | 6.48 (6.04, 6.90) | 3.16 (2.66, 3.69) | 6.20 (5.80, 6.59) | 3.64 (3.01, 4.32) |
| **Jan 20 - Jan 26** | 6.19 (5.77, 6.66) | 2.89 (2.37, 3.42) | 0.49 (0.27, 0.64) | 6.38 (5.97, 6.82) | 2.75 (2.25, 3.28) | 6.18 (5.79, 6.58) | 2.90 (2.40, 3.41) | 5.86 (5.50, 6.22) | 3.22 (2.65, 3.81) |
| **Jan 21 - Jan 27** | 5.82 (5.39, 6.32) | 2.50 (2.02, 3.06) | 0.44 (0.22, 0.62) | 5.99 (5.63, 6.41) | 2.41 (1.93, 2.95) | 5.77 (5.44, 6.16) | 2.54 (2.03, 3.08) | 5.40 (5.11, 5.76) | 2.71 (2.18, 3.34) |
| **Jan 22 - Jan 28** | 5.61 (5.25, 6.02) | 2.37 (1.89, 2.87) | 0.48 (0.29, 0.61) | 5.78 (5.43, 6.15) | 2.27 (1.83, 2.75) | 5.59 (5.26, 5.93) | 2.39 (1.91, 2.89) | 5.26 (4.97, 5.59) | 2.53 (1.99, 3.13) |
| **Jan 23 - Jan 29** | 5.49 (5.09, 5.89) | 2.91 (2.29, 3.43) | 0.31 (0.13, 0.47) | 5.61 (5.20, 6.00) | 2.85 (2.24, 3.42) | 5.40 (5.03, 5.77) | 2.97 (2.34, 3.57) | 5.06 (4.71, 5.42) | 3.19 (2.44, 3.94) |
| **Jan 24 - Jan 30** | 5.49 (5.10, 5.93) | 2.91 (2.29, 3.51) | 0.40 (0.22, 0.54) | 5.54 (5.19, 5.93) | 2.84 (2.29, 3.40) | 5.31 (4.96, 5.69) | 2.93 (2.34, 3.51) | 4.92 (4.60, 5.30) | 3.08 (2.30, 3.79) |
| **Jan 25 - Jan 31** | 5.30 (4.86, 5.75) | 3.01 (2.35, 3.56) | 0.38 (0.20, 0.52) | 5.43 (5.01, 5.83) | 3.00 (2.32, 3.55) | 5.16 (4.77, 5.55) | 3.04 (2.40, 3.59) | 4.75 (4.40, 5.12) | 3.12 (2.42, 3.75) |
| **Jan 26 - Feb 01** | 5.12 (4.69, 5.56) | 3.13 (2.54, 3.69) | 0.41 (0.25, 0.56) | 5.28 (4.87, 5.74) | 3.07 (2.48, 3.64) | 5.02 (4.62, 5.44) | 3.18 (2.61, 3.72) | 4.62 (4.23, 5.02) | 3.37 (2.66, 4.07) |
| **Jan 27 - Feb 02** | 5.21 (4.76, 5.68) | 3.39 (2.71, 4.04) | 0.47 (0.27, 0.63) | 5.40 (4.94, 5.90) | 3.23 (2.58, 3.84) | 5.19 (4.79, 5.66) | 3.42 (2.76, 4.03) | 4.88 (4.49, 5.30) | 3.88 (3.05, 4.66) |
| **Jan 28 - Feb 03** | 5.40 (4.92, 5.90) | 3.77 (2.98, 4.51) | 0.58 (0.43, 0.72) | 5.64 (5.14, 6.19) | 3.45 (2.71, 4.08) | 5.46 (5.00, 5.98) | 3.64 (2.87, 4.27) | 5.22 (4.79, 5.71) | 4.18 (3.23, 4.98) |
| **Jan 29 - Feb 04** | 5.32 (4.80, 5.83) | 3.95 (3.18, 4.72) | 0.61 (0.41, 0.76) | 5.62 (5.09, 6.19) | 3.59 (2.91, 4.29) | 5.41 (4.94, 5.93) | 3.77 (3.09, 4.46) | 5.16 (4.70, 5.64) | 4.33 (3.51, 5.28) |
| **Jan 30 - Feb 29** | 6.36 (5.74, 6.92) | 3.99 (3.11, 4.77) | 0.56 (0.35, 0.73) | 6.64 (5.97, 7.30) | 3.62 (2.75, 4.34) | 6.41 (5.78, 7.05) | 3.88 (2.99, 4.64) | 6.14 (5.57, 6.78) | 4.61 (3.44, 5.75) |

| | | | | Mean width of exposure window | | | | | | | | | |
|---|---|---|---|---|---|---|---|---|---|---|---|---|---|
| | | | | 1 day | | | | 4 days | | | | [illegible] | |
| GT setting | Time window | Mean of sample size | $\mu_{realized}^{emp}$ | $\mu_{realized}^{est}$ | Mean width of 95% CI | Prop. of 95% CI covered $\mu_{realized}^{emp}$ | Bias | $\mu_{realized}^{est}$ | Mean width of 95% CI | Prop. of 95% CI covered $\mu_{realized}^{emp}$ | Bias | $\mu_{realized}^{est}$ | Mea[illegible] 9[illegible] |
| **Long** | D10 - D19 | 76 | 9.49 | 9.47 | 2.59 | 98% | -0.13% | 9.46 | 2.64 | 98% | -0.30% | 9.53 | |
| | D20 - D29 | 163 | 9.04 | 8.97 | 1.70 | 92% | -0.82% | 8.93 | 1.73 | 92% | -1.25% | 8.98 | |
| | D30 - D39 | 239 | 8.62 | 8.58 | 1.28 | 88% | -0.48% | 8.53 | 1.31 | 86% | -1.01% | 8.52 | |
| | D40 - D49 | 209 | 8.44 | 8.51 | 1.39 | 90% | 0.76% | 8.46 | 1.44 | 88% | 0.18% | 8.35 | |
| | D50 - D59 | 109 | 8.68 | 8.70 | 2.07 | 96% | 0.22% | 8.62 | 2.14 | 94% | -0.72% | 8.43 | |
| | D60 - D100 | 57 | 9.28 | 9.25 | 3.16 | 98% | -0.37% | 9.17 | 3.27 | 98% | -1.22% | 8.98 | |
| **Medium** | D5 - D11 | 45 | 6.84 | 7.01 | 2.37 | 88% | 2.50% | 7.03 | 2.52 | 94% | 2.82% | 7.23 | |
| | D12 - D18 | 106 | 6.56 | 6.60 | 1.49 | 96% | 0.63% | 6.59 | 1.55 | 92% | 0.37% | 6.72 | |
| | D19 - D25 | 194 | 6.28 | 6.21 | 1.02 | 88% | -1.05% | 6.14 | 1.09 | 88% | -2.09% | 6.22 | |
| | D26 - D32 | 234 | 6.05 | 5.97 | 0.87 | 92% | -1.33% | 5.91 | 0.94 | 88% | -2.39% | 5.86 | |
| | D33 - D39 | 171 | 6.03 | 6.01 | 1.05 | 96% | -0.34% | 5.92 | 1.13 | 92% | -1.81% | 5.73 | |
| | D40 - D60 | 120 | 6.32 | 6.27 | 1.35 | 86% | -0.76% | 6.17 | 1.45 | 86% | -2.37% | 5.91 | |
| **Short** | D9 - D12 | 90 | 3.83 | 3.81 | 0.84 | 90% | -0.38% | 3.78 | 1.01 | 88% | -1.28% | 4.14 | |
| | D13 - D16 | 164 | 3.70 | 3.69 | 0.59 | 92% | -0.40% | 3.61 | 0.71 | 80% | -2.38% | 3.75 | |
| | D17 - D20 | 209 | 3.60 | 3.59 | 0.50 | 94% | -0.38% | 3.46 | 0.61 | 78% | -4.13% | 3.44 | |
| | D21 - D24 | 179 | 3.57 | 3.57 | 0.55 | 98% | 0.07% | 3.44 | 0.68 | 90% | -3.49% | 3.30 | |
| | D25 - D28 | 110 | 3.59 | 3.60 | 0.71 | 98% | 0.15% | 3.45 | 0.88 | 96% | -4.02% | 3.17 | |
| | D29 - D40 | 78 | 3.69 | 3.74 | 0.88 | 90% | 1.15% | 3.60 | 1.09 | 94% | -2.50% | 3.28 | |

| | | | | Mean width of exposure window | | | | | | | | |
|---|---|---|---|---|---|---|---|---|---|---|---|---|
| | | | | 1 day | | | | 4 days | | | | |
| GT setting | Time window | Mean of sample size | $\sigma_{realized}^{emp}$ | $\sigma_{realized}^{est}$ | Mean width of 95% CI | Prop. of 95% CI covered $\sigma_{realized}^{emp}$ | Bias | $\sigma_{realized}^{est}$ | Mean width of 95% CI | Prop. of 95% CI covered $\sigma_{realized}^{emp}$ | Bias | $\sigma_{realized}^{est}$ |
| **Long** | D10 - D19 | 76 | 5.45 | 5.48 | 2.34 | 94% | 0.54% | 5.41 | 2.49 | 92% | -0.60% | 5.70 |
| | D20 - D29 | 163 | 5.25 | 5.28 | 1.54 | 94% | 0.54% | 5.20 | 1.64 | 96% | -0.99% | 5.45 |
| | D30 - D39 | 239 | 5.06 | 5.01 | 1.15 | 98% | -1.06% | 4.93 | 1.23 | 100% | -2.58% | 5.23 |
| | D40 - D49 | 209 | 5.12 | 5.05 | 1.26 | 88% | -1.35% | 5.03 | 1.37 | 92% | -1.72% | 5.37 |
| | D50 - D59 | 109 | 5.33 | 5.32 | 1.90 | 92% | -0.29% | 5.29 | 2.06 | 92% | -0.75% | 5.63 |
| | D60 - D100 | 57 | 5.65 | 5.55 | 2.90 | 96% | -1.73% | 5.53 | 3.14 | 94% | -2.14% | 5.82 |
| **Medium** | D5 - D11 | 45 | 3.75 | 3.79 | 2.11 | 92% | 1.13% | 3.78 | 2.39 | 88% | 0.80% | 4.24 |
| | D12 - D18 | 106 | 3.62 | 3.68 | 1.33 | 96% | 1.49% | 3.57 | 1.46 | 96% | -1.40% | 3.88 |
| | D19 - D25 | 194 | 3.48 | 3.49 | 0.91 | 94% | 0.25% | 3.46 | 1.03 | 92% | -0.69% | 3.80 |
| | D26 - D32 | 234 | 3.43 | 3.39 | 0.79 | 92% | -1.04% | 3.35 | 0.89 | 86% | -2.25% | 3.65 |
| | D33 - D39 | 171 | 3.49 | 3.44 | 0.95 | 94% | -1.45% | 3.40 | 1.07 | 94% | -2.59% | 3.71 |
| | D40 - D60 | 120 | 3.69 | 3.61 | 1.22 | 86% | -2.24% | 3.59 | 1.39 | 88% | -2.92% | 3.87 |
| **Short** | D9 - D12 | 90 | 1.86 | 1.90 | 0.75 | 94% | 2.19% | 1.87 | 0.96 | 98% | 0.83% | 2.29 |
| | D13 - D16 | 164 | 1.80 | 1.82 | 0.52 | 98% | 0.98% | 1.77 | 0.67 | 92% | -1.85% | 2.12 |
| | D17 - D20 | 209 | 1.78 | 1.80 | 0.44 | 92% | 1.36% | 1.77 | 0.57 | 96% | -0.36% | 2.04 |
| | D21 - D24 | 179 | 1.78 | 1.80 | 0.48 | 92% | 0.70% | 1.76 | 0.63 | 98% | -1.20% | 1.99 |
| | D25 - D28 | 110 | 1.81 | 1.82 | 0.63 | 98% | 0.15% | 1.80 | 0.82 | 98% | -1.01% | 1.95 |
| | D29 - D40 | 78 | 1.87 | 1.91 | 0.78 | 92% | 2.36% | 1.90 | 1.03 | 96% | 1.75% | 2.05 |

| | | | | Mean width of exposure windo | | | | | | |
|---|---|---|---|---|---|---|---|---|---|---|
| | | | | 1 day | | | | 4 days | | |
| **GT setting** | **Time window** | **Mean of sample size** | $\mu^{emp}_{realized}$ | $\mu^{est}_{realized}$ | **Mean width of 95% CI** | **Prop. of 95% CI covered $\mu^{emp}_{realized}$** | **Bias** | $\mu^{est}_{realized}$ | **Mean width of 95% CI** | **Prop. of 95% covered $\mu^{emp}_{rea}$** |
| **Medium** | D5 - D11 | 45 | 6.84 | 6.98 | 2.50 | 84% | 2.03% | 6.94 | 2.65 | 94% |
| | D12 - D18 | 106 | 6.56 | 6.44 | 1.82 | 86% | -1.78% | 6.40 | 1.85 | 72% |
| | D19 - D25 | 194 | 6.28 | 6.07 | 1.25 | 78% | -3.29% | 5.96 | 1.29 | 78% |
| | D26 - D32 | 234 | 6.05 | 5.78 | 1.09 | 74% | -4.57% | 5.61 | 1.14 | 68% |
| | D33 - D39 | 171 | 6.03 | 5.97 | 1.36 | 92% | -0.85% | 5.80 | 1.41 | 84% |
| | D40 - D60 | 120 | 6.32 | 6.20 | 1.80 | 88% | -1.89% | 6.00 | 1.87 | 84% |

**Supplementary Table 8. Comparison between the estimated standard deviation (SD) of forward generation time (GT) and the SD of realized GT i**

Medium GT setting was used to assess the recovery performance of model with a mean of 7 days and a SD of 4 days. $\sigma^{emp}_{realized}$: mean of empirical SD of

$\sigma^{est}_{realized}/\sigma^{emp}_{realized} - 1$. Recovery performance was also assessed with different mean widths of exposure window (1, 4, and 7 days respectively) for remaini

| | | | | Mean width of exposure windo | | | | | | |
|---|---|---|---|---|---|---|---|---|---|---|
| | | | | 1 day | | | | 4 days | | |
| **GT setting** | **Time window** | **Mean of sample size** | $\sigma^{emp}_{realized}$ | $\sigma^{est}_{realized}$ | **Mean width of 95% CI** | **Prop. of 95% CI covered $\sigma^{emp}_{realized}$** | **Bias** | $\sigma^{est}_{realized}$ | **Mean width of 95% CI** | **Prop. of 95% covered $\sigma^{emp}_{reali}$** |
| **Medium** | D5 - D11 | 45 | 3.75 | 3.80 | 2.29 | 96% | 1.56% | 3.86 | 2.58 | 94% |
| | D12 - D18 | 106 | 3.62 | 3.72 | 1.77 | 88% | 2.75% | 3.70 | 1.85 | 90% |
| | D19 - D25 | 194 | 3.48 | 3.53 | 1.21 | 86% | 1.37% | 3.56 | 1.30 | 88% |
| | D26 - D32 | 234 | 3.43 | 3.45 | 1.07 | 94% | 0.69% | 3.49 | 1.14 | 96% |
| | D33 - D39 | 171 | [illegible] | 3.61 | 1.34 | 92% | 3.37% | 3.64 | 1.43 | 92% |

EpiEstim package[4]. $\mu_{realized}^{emp}$: mean of empirical mean of realized GT; $\sigma_{realized}^{emp}$: mean of empirical SD of realized GT; ${R_I}_{realized}^{emp}$: mean of realized initial eff

| GT setting | Time window | Mean of sample size | $\mu_{realized}^{emp}$ | $\sigma_{realized}^{emp}$ | ${R_I}_{realized}^{emp}$ | Distribution used for $R_I$ calculation | Mean width of exposure windows | Mean | | Standard devia | |
|---|---|---|---|---|---|---|---|---|---|---|---|
| | | | | | | | | **Estimate** | **Bias** | **Estimate** | **Bi** |
| **Long** | D1 - D15 | 66 | 9.70 | 5.59 | 2.35 | SI | - | 10.92 | 12.55% | 7.02 | 25.7 |
| | D1 - D15 | 66 | 9.70 | 5.59 | 2.35 | GT | 1 day | 9.61 | -0.99% | 5.64 | 1.0 |
| | D1 - D15 | 66 | 9.70 | 5.59 | 2.35 | GT | 4 days | 9.62 | -0.88% | 5.63 | 0.7 |
| | D1 - D15 | 66 | 9.70 | 5.59 | 2.35 | GT | 7 days | 9.83 | 1.29% | 5.92 | 5.9 |
| | D1 - D15 | 66 | 9.70 | 5.59 | 2.35 | GT | 10 days | 9.99 | 2.91% | 6.29 | 12.6 |
| **Medium** | D1 - D10 | 45 | 6.88 | 3.77 | 2.37 | SI | - | 8.96 | 30.30% | 5.46 | 44.9 |
| | D1 - D10 | 45 | 6.88 | 3.77 | 2.37 | GT | 1 day | 6.98 | 1.47% | 3.81 | 1.1 |
| | D1 - D10 | 45 | 6.88 | 3.77 | 2.37 | GT | 4 days | 6.98 | 1.53% | 3.79 | 0.5 |
| | D1 - D10 | 45 | 6.88 | 3.77 | 2.37 | GT | 7 days | 7.15 | 4.01% | 4.21 | 11.7 |
| | D1 - D10 | 45 | 6.88 | 3.77 | 2.37 | GT | 10 days | 7.33 | 6.57% | 4.51 | 19.5 |
| **Short** | D1 - D10 | 85 | 3.91 | 1.91 | 2.13 | SI | - | 6.24 | 59.51% | 4.43 | 132. |
| | D1 - D10 | 85 | 3.91 | 1.91 | 2.13 | GT | 1 day | 3.89 | -0.47% | 1.94 | 1.8 |
| | D1 - D10 | 85 | 3.91 | 1.91 | 2.13 | GT | 4 days | 3.94 | 0.68% | 1.94 | 1.6 |
| | D1 - D10 | 85 | 3.91 | 1.91 | 2.13 | GT | 7 days | 4.25 | 8.63% | 2.27 | 19.2 |
| | D1 - D10 | 85 | 3.91 | 1.91 | 2.13 | GT | 10 days | 4.34 | 10.89% | 2.59 | 35.8 |

estimated mean of realized GT; CI: confidence interval; Bias: $\mu^{est}_{realized}/\mu^{emp}_{realized} - 1$.

| | | | | | | | | | | |
|---|---|---|---|---|---|---|---|---|---|---|
| | | | | **Mean width of exposure windows** | | | | | | |
| | | | | **1 day** | | | | **4 days** | | |
| **Correlation setting** | **Time window** | **Mean of sample size** | $\mu^{emp}_{realized}$ | $\mu^{est}_{realized}$ | **Mean width of 95% CI** | **Prop. of 95% CI covered $\mu^{emp}_{realized}$** | **Bias** | $\mu^{est}_{realized}$ | **Mean width of 95% CI** | **Prop. covere** |
| $\rho = 0$ | D5 - D10 | 29 | 6.75 | 6.72 | 2.45 | 92% | -0.38% | 7.17 | 2.94 | |
| | D10 – D15 | 54 | 6.65 | 6.68 | 1.77 | 94% | 0.40% | 7.04 | 1.84 | |
| | D15 – D20 | 95 | 6.46 | 6.48 | 1.31 | 96% | 0.19% | 6.85 | 1.37 | |
| | D20 – D25 | 141 | 6.27 | 6.37 | 1.04 | 94% | 1.61% | 6.66 | 1.08 | |
| | D25 – D30 | 169 | 6.11 | 6.18 | 0.90 | 94% | 1.13% | 6.52 | 0.94 | |
| | D30 – D35 | 156 | 6.08 | 6.12 | 0.93 | 94% | 0.69% | 6.52 | 0.96 | |
| $\rho = 0.25$ | D5 - D10 | 29 | 6.48 | 6.25 | 2.16 | 82% | -3.52% | 6.62 | 2.40 | |
| | D10 – D15 | 55 | 6.42 | 6.34 | 1.60 | 96% | -1.28% | 6.61 | 1.60 | |
| | D15 – D20 | 97 | 6.27 | 6.21 | 1.20 | 98% | -0.93% | 6.48 | 1.23 | |
| | D20 – D25 | 143 | 6.13 | 6.23 | 0.96 | 88% | 1.63% | 6.43 | 0.99 | |
| | D25 – D30 | 171 | 6.05 | 6.13 | 0.87 | 88% | 1.26% | 6.37 | 0.86 | |
| | D30 – D35 | 156 | 6.12 | 6.14 | 0.90 | 96% | 0.25% | 6.42 | 0.92 | |
| $\rho = 0.50$ | D5 - D10 | 29 | 6.17 | 6.07 | 0.90 | 82% | 0.25% | 6.26 | 2.15 | |
| | D10 – D15 | 55 | 6.17 | 6.12 | 1.89 | 92% | -1.62% | 6.40 | 1.37 | |
| | D15 – D20 | 98 | 6.06 | 6.07 | 1.42 | 86% | -0.84% | 6.29 | 1.06 | |
| | D20 – D25 | 145 | 6.00 | 6.11 | 1.07 | 86% | 0.13% | 6.29 | 0.83 | |
| | D25 – D30 | 173 | 6.02 | 6.05 | 0.87 | 84% | 1.84% | 6.24 | 0.74 | |
| | D30 – D35 | 155 | 6.19 | 6.18 | 0.77 | 88% | 0.51% | 6.41 | 0.83 | |
| $\rho = 0.75$ | D5 - D10 | 29 | 5.84 | 5.74 | 1.42 | 74% | -1.76% | 5.91 | 1.60 | |
| | D10 – D15 | 55 | 5.93 | 5.87 | 1.06 | 78% | -0.99% | 6.04 | 1.09 | |
| | D15 – D20 | 99 | 5.88 | 5.87 | 0.81 | 58% | -0.17% | 6.00 | 0.84 | |
| | D20 – D25 | 147 | 5.88 | 5.94 | 0.66 | 50% | 1.02% | 6.03 | 0.66 | |
| | D25 – D30 | 174 | 5.98 | 6.02 | 0.61 | 66% | 0.65% | 6.16 | 0.63 | |
| | D30 – D35 | 155 | 6.26 | 6.22 | 0.66 | 66% | -0.70% | 6.38 | 0.66 | |

$\sigma^{est}_{realized}$: mean of estimated SD of realized GT; CI: confidence interval; Bias: $\sigma^{est}_{realized}/\sigma^{emp}_{realized} - 1$.

| | | | | Mean width of exposure windows | | | | | | |
|---|---|---|---|---|---|---|---|---|---|---|
| | | | | 1 day | | | | 4 days | | |
| **Correlation setting** | **Time window** | **Mean of sample size** | $\sigma^{emp}_{realized}$ | $\sigma^{est}_{realized}$ | **Mean width of 95% CI** | **Prop. of 95% CI covered $\sigma^{emp}_{realized}$** | **Bias** | $\sigma^{est}_{realized}$ | **Mean width of 95% CI** | **Prop. o[illegible] covere[illegible]** |
| $\rho = 0$ | D5 - D10 | 29 | 3.51 | 3.48 | 2.76 | 96% | -0.77% | 3.64 | 3.55 | 9[illegible] |
| | D10 – D15 | 54 | 3.48 | 3.53 | 2.01 | 100% | 1.54% | 3.38 | 2.15 | 9[illegible] |
| | D15 – D20 | 95 | 3.36 | 3.43 | 1.48 | 98% | 2.17% | 3.25 | 1.52 | 9[illegible] |
| | D20 – D25 | 141 | 3.23 | 3.36 | 1.17 | 96% | 3.93% | 3.08 | 1.14 | 8[illegible] |
| | D25 – D30 | 169 | 3.18 | 3.23 | 1.01 | 96% | 1.47% | 2.95 | 1.04 | 8[illegible] |
| | D30 – D35 | 156 | 3.25 | 3.22 | 1.04 | 94% | -0.78% | 3.02 | 1.04 | 9[illegible] |
| $\rho = 0.25$ | D5 - D10 | 29 | 3.38 | 3.22 | 2.48 | 88% | -4.78% | 3.31 | 3.03 | 9[illegible] |
| | D10 – D15 | 55 | 3.38 | 3.27 | 1.82 | 94% | -3.06% | 2.97 | 1.85 | 8[illegible] |
| | D15 – D20 | 97 | 3.26 | 3.26 | 1.37 | 96% | -0.09% | 2.97 | 1.38 | 8[illegible] |
| | D20 – D25 | 143 | 3.17 | 3.26 | 1.10 | 100% | 2.69% | 2.90 | 1.11 | 8[illegible] |
| | D25 – D30 | 171 | 3.15 | 3.20 | 0.99 | 96% | 1.58% | 2.89 | 0.94 | 7[illegible] |
| | D30 – D35 | 156 | 3.23 | 3.22 | 1.03 | 96% | -0.28% | 2.96 | 1.01 | 7[illegible] |
| $\rho = 0.50$ | D5 - D10 | 29 | 3.11 | 3.02 | 2.22 | 88% | -2.99% | 2.95 | 2.82 | 8[illegible] |
| | D10 – D15 | 55 | 3.20 | 3.17 | 1.69 | 96% | -1.03% | 2.77 | 1.74 | 7[illegible] |
| | D15 – D20 | 98 | 3.14 | 3.16 | 1.28 | 98% | 0.60% | 2.82 | 1.31 | 7[illegible] |
| | D20 – D25 | 145 | 3.08 | 3.19 | 1.05 | 94% | 3.46% | 2.78 | 1.05 | 7[illegible] |
| | D25 – D30 | 173 | 3.11 | 3.13 | 0.93 | 96% | 0.52% | 2.77 | 0.92 | 6[illegible] |
| | D30 – D35 | 155 | 3.21 | 3.23 | 1.00 | 94% | 0.60% | 2.96 | 1.04 | 7[illegible] |
| $\rho = 0.75$ | D5 - D10 | 29 | 2.72 | 2.74 | 1.77 | 84% | 0.50% | 2.61 | 2.41 | 9[illegible] |
| | D10 – D15 | 55 | 2.98 | 2.92 | 1.35 | 90% | -1.94% | 2.66 | 1.64 | 7[illegible] |
| | D15 – D20 | 99 | 2.99 | 2.97 | 1.04 | 80% | -0.69% | 2.72 | 1.20 | 7[illegible] |
| | D20 – D25 | 147 | 2.98 | 3.03 | 0.85 | 78% | 1.69% | 2.76 | 0.91 | 6[illegible] |
| | D25 – D30 | 174 | 3.03 | 3.09 | 0.79 | 72% | 1.76% | 2.77 | 0.89 | 7[illegible] |
| | D30 – D35 | 155 | 3.18 | 3.19 | 0.85 | 76% | 0.19% | 2.89 | 0.90 | 6[illegible] |

$\rho = 0$ was not reported given the small magnitude of $\tilde{\rho}^{emp}_{realized}$.

| | | | | Mean width of exposure windows | | | | | | |
|---|---|---|---|---|---|---|---|---|---|---|
| | | | | 1 day | | | | 4 days | | |
| **Correlation setting** | **Time window** | **Mean of sample size** | $\tilde{\boldsymbol{\rho}}^{emp}_{realized}$ | $\tilde{\boldsymbol{\rho}}^{est}_{realized}$ | **Mean width of 95% CI** | **Prop. of 95% CI covered** $\tilde{\boldsymbol{\rho}}^{emp}_{realized}$ | **Bias** | $\tilde{\boldsymbol{\rho}}^{est}_{realized}$ | **Mean width of 95% CI** | **Prop[illegible] cove[illegible]** |
| $\boldsymbol{\rho = 0}$* | D5 - D10 | 29 | 0.01 | -0.01 | 0.71 | 92% | - | -0.24 | 0.97 | |
| | D10 – D15 | 54 | 0.02 | -0.02 | 0.51 | 82% | - | -0.19 | 0.69 | |
| | D15 – D20 | 95 | 0.02 | -0.01 | 0.39 | 84% | - | -0.14 | 0.52 | |
| | D20 – D25 | 141 | 0.03 | 0.02 | 0.32 | 96% | - | -0.12 | 0.44 | |
| | D25 – D30 | 169 | 0.03 | 0.01 | 0.29 | 96% | - | -0.11 | 0.39 | |
| | D30 – D35 | 156 | 0.01 | 0.00 | 0.30 | 100% | - | -0.14 | 0.39 | |
| $\boldsymbol{\rho = 0.25}$ | D5 - D10 | 29 | 0.19 | 0.20 | 0.66 | 92% | 2.88% | 0.02 | 0.96 | |
| | D10 – D15 | 55 | 0.22 | 0.19 | 0.49 | 88% | -10.49% | 0.05 | 0.68 | |
| | D15 – D20 | 97 | 0.22 | 0.21 | 0.37 | 96% | -2.95% | 0.12 | 0.54 | |
| | D20 – D25 | 143 | 0.23 | 0.24 | 0.29 | 88% | 4.10% | 0.13 | 0.43 | |
| | D25 – D30 | 171 | 0.23 | 0.23 | 0.27 | 94% | -0.09% | 0.14 | 0.38 | |
| | D30 – D35 | 156 | 0.21 | 0.23 | 0.28 | 96% | 9.24% | 0.15 | 0.37 | |
| $\boldsymbol{\rho = 0.50}$ | D5 - D10 | 29 | 0.38 | 0.39 | 0.57 | 92% | 0.99% | 0.20 | 0.96 | |
| | D10 – D15 | 55 | 0.42 | 0.42 | 0.40 | 78% | -1.83% | 0.33 | 0.58 | |
| | D15 – D20 | 98 | 0.44 | 0.45 | 0.29 | 88% | 2.89% | 0.38 | 0.44 | |
| | D20 – D25 | 145 | 0.45 | 0.46 | 0.24 | 90% | 3.49% | 0.39 | 0.36 | |
| | D25 – D30 | 173 | 0.44 | 0.47 | 0.21 | 86% | 4.85% | 0.42 | 0.30 | |
| | D30 – D35 | 155 | 0.44 | 0.46 | 0.22 | 86% | 4.80% | 0.40 | 0.31 | |
| $\boldsymbol{\rho = 0.75}$ | D5 - D10 | 29 | 0.62 | 0.65 | 0.38 | 76% | 6.02% | 0.58 | 0.62 | |
| | D10 – D15 | 55 | 0.67 | 0.68 | 0.24 | 82% | 1.45% | 0.65 | 0.40 | |
| | D15 – D20 | 99 | 0.68 | 0.69 | 0.18 | 80% | 1.64% | 0.68 | 0.27 | |
| | D20 – D25 | 147 | 0.69 | 0.71 | 0.14 | 86% | 2.14% | 0.70 | 0.19 | |
| | D25 – D30 | 174 | 0.69 | 0.71 | 0.13 | 88% | 2.37% | 0.71 | 0.17 | |
| | D30 – D35 | 155 | 0.69 | 0.71 | 0.13 | 76% | 3.62% | 0.70 | 0.17 | |
